# Identifying protein biomarkers and therapeutic targets in psoriasis through integrative genomic, proteomic and transcriptomic analysis

**DOI:** 10.64898/2026.07.09.26357649

**Authors:** Devendra Meena, Christos V. Chalitsios, Jingxian Huang, Narendra Meena, Siwei Wu, Alexander Smith, Charalabos Antonatos, Yiannis Vasilopoulos, James Yarmolinsky, Dipender Gill, Abbas Dehghan, Konstantinos K. Tsilidis, Ioanna Tzoulaki

## Abstract

Plasma proteins are promising biomarkers and potential drug targets in psoriasis. We conducted a two-sample Mendelian randomisation analysis integrating protein quantitative trait loci from UK Biobank and deCODE genetics with a psoriasis GWAS meta-analysis of 36,466 cases. To strengthen causal inference, we performed colocalisation analyses to evaluate shared genetic signals and applied summary data–based MR (SMR) with HEIDI testing using expression quantitative trait loci to exclude linkage-driven associations. After correction for multiple testing, 78 circulating proteins showed genetically predicted associations with psoriasis, with 27 demonstrating strong colocalisation (PPH4>80%). Triangulation prioritised 12 Tier 1 proteins, STX4, FLT3, NFKB1, IL18, PRSS53, SPAG1, SGSH, PLAT, RALB, TNFSF11, SPHK2, and STAT3, supported by consistent effects and no heterogeneity. Network profiling and Genome for REPositioning analyses assessed biological connectivity and druggability, revealing enrichment in anatomical therapeutic chemical groups L and B. Single-cell RNA sequencing confirmed cell-type–specific expression and modulation following IL-23 blockade.

## INTRODUCTION

Psoriasis is a chronic, immune-mediated inflammatory skin disorder affecting over 125 million individuals globally (1–3). It manifests with a heterogeneous clinical spectrum of cutaneous and systemic features (3), with plaque psoriasis (psoriasis vulgaris) representing the most prevalent subtype, accounting for approximately 80% of cases (2). Patients often experience itching, burning sensations, and skin discomfort, leading to substantial impairments in daily functioning and quality of life (4). Despite its considerable individual and societal burden, the pathogenesis of psoriasis remains incompletely understood, hindering the development of curative or universally effective therapies.

Current evidence suggests that psoriasis arises from a complex interplay between genetic predisposition and environmental triggers, initiating the activation of plasmacytoid dendritic cells and a downstream immune cascade (5). This leads to the overproduction of key proinflammatory cytokines, including tumour necrosis factor (TNF)-α, interferon-gamma (IFN-γ), and interleukins such as IL-1β, IL-17, IL-22, and IL-23 (5). These cytokines drive keratinocyte hyperproliferation and aberrant differentiation, thereby sustaining a self-amplifying inflammatory loop (6). Advances in this mechanistic understanding have transformed psoriasis management with the introduction of biologic therapies targeting TNF-α, IL-17, and IL-23 (7). Nonetheless, treatment responses remain heterogeneous, and a substantial proportion of patients experience inadequate efficacy or adverse effects, highlighting the need for novel and more precisely targeted treatment strategies (7,8).

Plasma proteins play central roles in both physiological regulation and disease pathogenesis (9–12) and constitute a major class of druggable targets, with a large proportion of recently FDA-approved drugs acting on human proteins (13,14). Genome-wide association studies (GWAS) of circulating protein levels have identified genetic variants associated with protein abundance, known as protein quantitative trait loci (pQTLs) (15–18). These pQTLs enable the application of Mendelian randomisation (MR) to infer the potential causal role of proteins in disease pathogenesis, leveraging the quasi-random allocation of alleles at conception to minimise confounding and reverse causation (19–21).

Prior proteome-wide MR studies of psoriasis have been limited by smaller outcome GWAS datasets, single-platform proteomic coverage, or limited colocalisation and prioritisation frameworks (22,23). In this study, we address these gaps by leveraging the largest available psoriasis GWAS, integrating complementary large-scale proteomic platforms, and applying a multi-layer prioritisation framework incorporating pairwise conditional colocalisation (PWCoCo), tissue-specific summary-data–based Mendelian randomisation (SMR) coupled with heterogeneity in dependent instruments (HEIDI), and network-based functional profiling to enhance robustness and biological interpretability. To further strengthen biological relevance, we performed independent single-cell RNA sequencing (scRNA-seq) analyses of healthy, non-lesional, and lesional psoriatic skin, and evaluated transcriptional changes following IL-23 blockade (risankizumab) using longitudinal lesional scRNA-seq datasets (**Figure 1**).

**Figure 1.**
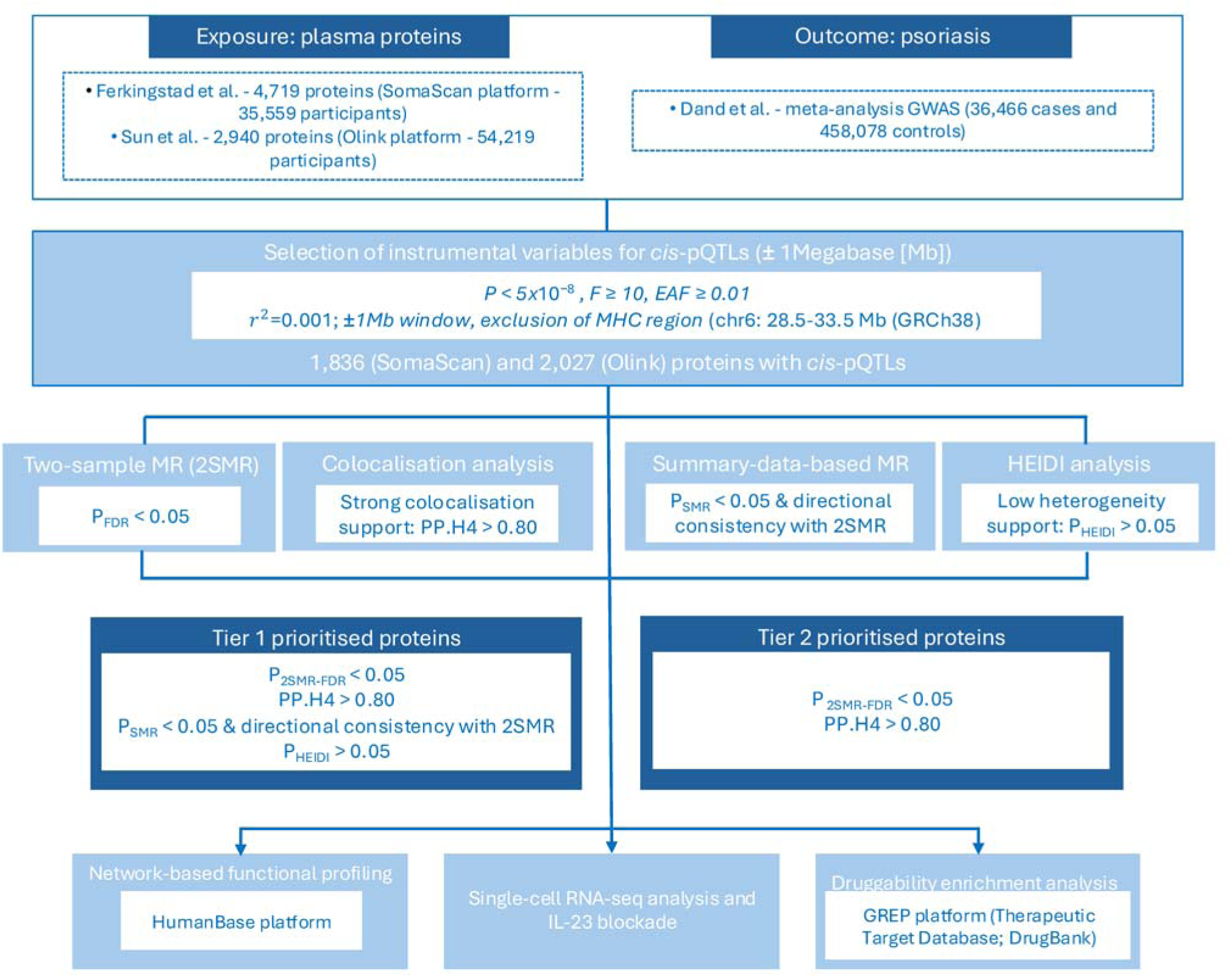
Flowchart of the study design.

## RESULTS

### Two-sample Mendelian randomisation and colocalisation analysis

The genetic instrument selection strategy enabled the evaluation of 2,027 and 1,836 plasma proteins with *cis*-pQTLs derived from the UK Biobank Pharma Proteomics Project (UKB-PPP) and deCODE genetics, respectively. All selected instruments exhibited F-statistics greater than 10, confirming sufficient instrument strength. Applying either the Wald ratio or inverse variance weighted (IVW) method, 51 proteins in UKB-PPP and 33 proteins in deCODE genetics were significantly associated with psoriasis risk after correction for multiple testing (P_2SMR_-_FDR_< 0.05), representing 78 unique proteins across the two datasets (**Table S1-S2, Figure 2A-2B**). Among these, genetically predicted higher plasma levels (per 1-SD) of 33 proteins were associated with a higher risk of psoriasis, while higher levels of 45 plasma proteins were associated with a lower risk. These associations were generally consistent across MR sensitivity analyses, including MR-Egger and the weighted median, with no evidence of heterogeneity (P_Cochran’s Q statistic_>0.05) or directional pleiotropy (P_MR-Egger intercept_>0.05).

**Figure 2.**
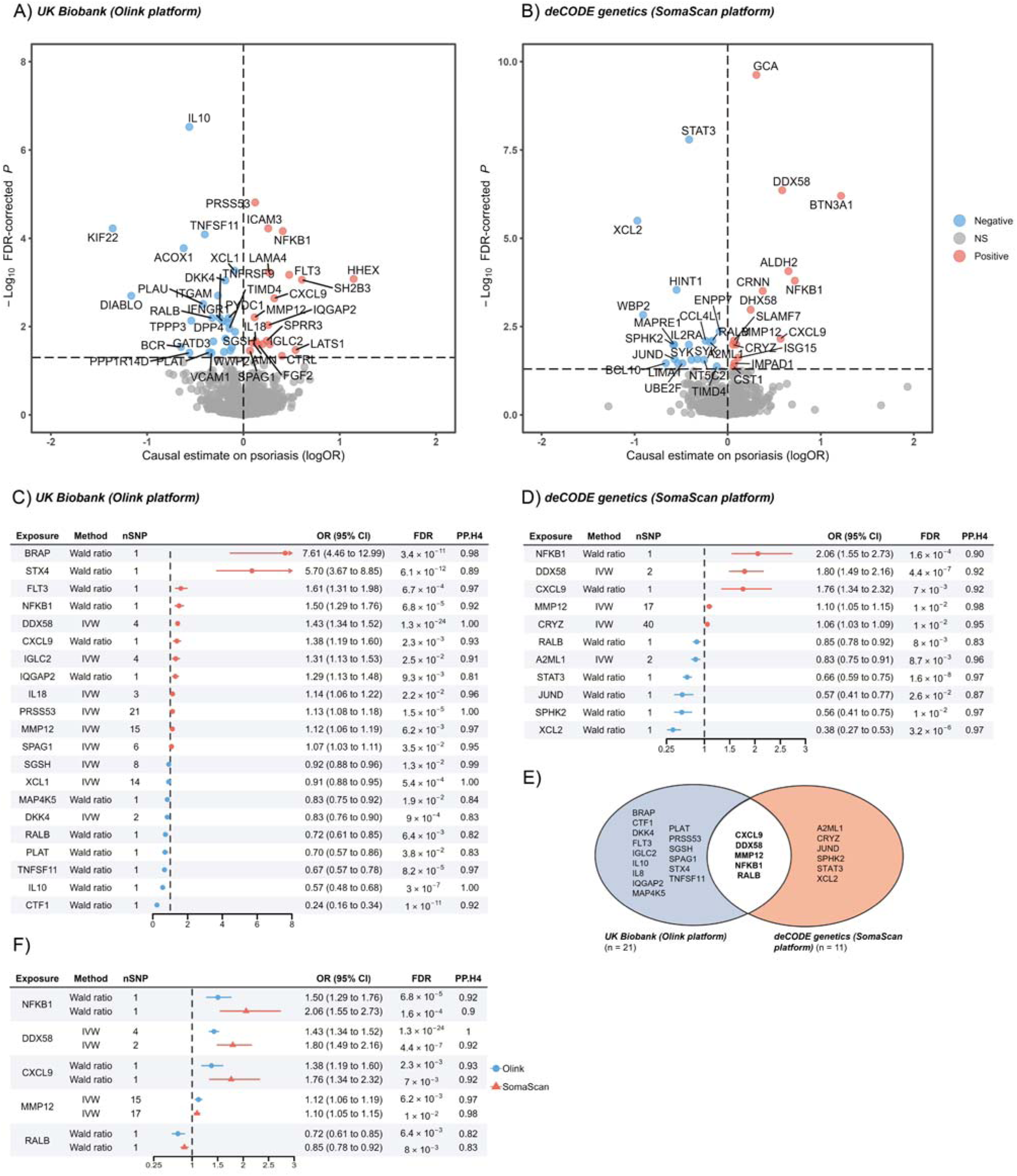
Volcano plots of two-sample MR results for (A) 2,027 *cis*-pQTLs of plasma proteins from the UK Biobank Pharma Proteomics Project (Olink platform) and (B) 1,836 *cis*-pQTLs from plasma proteins from deCODE genetics (Somascan platform) on the risk of psoriasis. Labelled proteins tested with significant P_2SMR_-_FDR_<0.05. (C) Forest plots displaying odds ratios (OR) and 95% confidence intervals for proteins significantly associated with psoriasis in MR analyses using UK Biobank (Olink platform) and (D) deCODE (SomaScan platform) data. OR for the risk of psoriasis is expressed as per 1-SD increase in plasma protein level. The posterior probability (PP.H4) of colocalisation is also shown. (E) Venn diagram summarising proteins with MR and colocalisation evidence from both platforms. CI, Confidence Interval; FDR, False Discovery Rate; IVW, Inverse Variance Weighted; OR, Odds Ratio; SNP, Single Nucleotide Polymorphism.

Of the proteins that showed evidence for a genetic association with psoriasis (P_2SMR_-_FDR_<0.05), 21 in the UKB-PPP and 11 in deCODE genetics (27 unique proteins) were further supported by strong colocalisation evidence (posterior probability pf hypotheses 4 [PP.H4]>0.80), indicating a high likelihood that the same genetic variant underlies both protein abundance and psoriasis susceptibility (**Figure 2C-2D, Table S3, Figures S1-S32**). Notably, five proteins (CXCL9, DDX58, MMP12, NFKB1, and RALB) were consistently identified across both platforms (**Figure 2E-2F**).

### Summary-based MR and HEIDI analysis

From the 27 unique proteins identified from the two-sample MR with strong colocalisation support (PP.H4>0.80), 12 genes encoding the above proteins also satisfied multiple lines of integrative evidence. Specifically, SMR analysis provided additional supportive evidence (P_SMR_<0.05), exhibited consistent effect direction with the primary two-sample MR estimates, and showed no evidence of heterogeneity based on the HEIDI analysis (P_HEIDI_>0.05) (**Table 1**). In psoriasis-relevant tissues, elevated genetically predicted transcription levels of *STX4, FLT3, NFKB1, IL18, PRSS53, SPAG1, SGSH,* and *PLAT* were associated with a higher risk of psoriasis. In contrast, higher genetically predicted expression of *RALB, TNFSF11, SPHK2, and STAT3* was associated with a lower risk of psoriasis.

**Table 1.** Summary data–based Mendelian randomisation (SMR) and HEIDI analyses results for genes encoding proteins. Included are proteins identified

| Gene | topSNP | topSNP<sub>chr</sub> | topSNP<sub>bp</sub> | P<sub>GWAS</sub><sup>a</sup> | P<sub>eQTL</sub><sup>b</sup> | B<sub>SMR</sub> | P<sub>SMR</sub><sup>c</sup> | P<sub>HEIDI</sub><sup>d</sup> | nSNP<sub>HEIDI</sub> | Tissue |
| --- | --- | --- | --- | --- | --- | --- | --- | --- | --- | --- |
| STX4 | rs58726213 | 16 | 31044683 | 1.50E-14 | 2.83E-08 | 0.22 | 6.64E-06 | 4.42E-01 | 4 | Cells EBV-transformed lymphocytes |
| STX4 | rs17839567 | 16 | 31057945 | 6.58E-15 | 9.72E-10 | 0.48 | 1.50E-06 | 3.68E-01 | 7 | Esophagus Mucosa |
| STX4 | rs7203999 | 16 | 31017554 | 7.38E-15 | 3.68E-12 | 0.27 | 2.06E-07 | 1.03E-01 | 9 | Spleen |
| FLT3 | rs12430881 | 13 | 28594802 | 2.59E-02 | 1.71E-11 | 0.11 | 3.44E-02 | 1.16E-01 | 10 | Esophagus Mucosa |
| FLT3 | rs61944200 | 13 | 28602256 | 4.86E-03 | 4.94E-15 | 0.10 | 7.91E-03 | 5.66E-02 | 15 | Skin Not Sun Exposed |
| FLT3 | rs61944199 | 13 | 28601651 | 5.10E-03 | 2.17E-08 | 0.08 | 1.21E-02 | 1.28E-01 | 6 | Spleen |
| NFKB1 | rs230520 | 4 | 103465612 | 5.28E-07 | 1.49E-12 | 0.26 | 4.55E-05 | 1.45E-01 | 13 | Fibroblast |
| IL18 | rs5744222 | 11 | 112037014 | 1.74E-03 | 7.83E-11 | 0.21 | 4.90E-03 | 2.72E-01 | 7 | Esophagus Mucosa |
| PRSS53 | rs59061704 | 16 | 31062704 | 1.50E-14 | 1.55E-14 | 0.30 | 5.49E-08 | 8.42E-02 | 15 | Skin Sun Exposed |
| SPAG1 | rs1788198 | 8 | 101292360 | 7.52E-06 | 1.54E-08 | 0.34 | 4.33E-04 | 6.23E-02 | 7 | Skin Not Sun Exposed |
| SPAG1 | rs1788190 | 8 | 101253184 | 8.54E-06 | 2.94E-09 | 0.41 | 3.61E-04 | 8.55E-02 | 9 | Skin Sun Exposed |
| SGSH | rs6565649 | 17 | 78192414 | 1.51E-11 | 4.29E-08 | 0.18 | 2.07E-05 | 2.33E-01 | 6 | Cells EBV-transformed lymphocytes |
| SGSH | rs9904453 | 17 | 78176579 | 8.58E-17 | 6.34E-23 | 0.26 | 2.23E-10 | 1.09E-01 | 20 | Esophagus Mucosa |
| SGSH | rs9904453 | 17 | 78176579 | 8.58E-17 | 3.18E-25 | 0.25 | 9.36E-11 | 2.38E-01 | 20 | Skin Not Sun Exposed |
| SGSH | rs9904453 | 17 | 78176579 | 8.58E-17 | 2.57E-25 | 0.33 | 9.06E-11 | 6.89E-02 | 20 | Skin Sun Exposed |
| SGSH | rs9904453 | 17 | 78176579 | 8.58E-17 | 2.14E-16 | 0.20 | 5.38E-09 | 2.91E-01 | 13 | Spleen |
| SGSH | rs8078181 | 17 | 78181427 | 2.71E-02 | 4.93E-08 | 0.12 | 4.04E-02 | 5.47E-02 | 4 | Stomach |
| RALB | rs3980213 | 2 | 121004979 | 8.34E-05 | 2.80E-14 | -0.05 | 4.81E-04 | 1.37E-01 | 11 | Cells EBV-transformed lymphocytes |
| RALB | rs12711940 | 2 | 121036386 | 1.05E-04 | 2.00E-11 | -0.20 | 7.52E-04 | 5.51E-01 | 10 | Skin Sun Exposed |
| PLAT | rs2020920 | 8 | 42064994 | 3.00E-05 | 2.69E-20 | 0.20 | 1.46E-04 | 1.67E-01 | 20 | Esophagus Mucosa |
| TNFSF11 | rs35489299 | 13 | 43085060 | 3.88E-03 | 2.66E-08 | -0.09 | 1.06E-02 | 7.95E-02 | 6 | Esophagus Mucosa |
| SPHK2 | rs443527 | 19 | 49116760 | 9.77E-04 | 4.49E-25 | -0.10 | 1.66E-03 | 4.57E-01 | 15 | Esophagus Mucosa |
| STAT3 | rs3024971 | 12 | 57493727 | 5.93E-03 | 1.57E-08 | -0.27 | 1.31E-02 | 4.41E-01 | 4 | Fibroblast |

<sup>a</sup> P<sub>GWAS</sub>: p-value for the top-associated eQTL in the psoriasis GWAS.
<sup>b</sup> P<sub>eQTL</sub>: P-value for the top associated eQTL in the GTEx data.
<sup>c</sup> P<sub>SMR</sub> is the p-value for SMR analysis
<sup>d</sup> P<sub>HEIDI</sub> is the p-value for the HEIDI analysis.

### Prioritised proteins

Integrating the lines of evidence described above, we categorised the proteins into two prioritisation tiers. Tier 1 comprised twelve proteins (STX4, FLT3, NFKB1, IL18, PRSS53, SPAG1, SGSH, PLAT, RALB, TNFSF11, SPHK2, and STAT3) which passed all prioritisation criteria, suggesting robust evidence of genetic association. Tier 2 included fifteen proteins (BRAP, DDX58, CXCL9, IGLC2, MMP12, XCL1, MAP4K5, DKK4, IL10, CTF1, CRYZ, A2ML1, JUND, and XCL2), which failed one or both of the SMR and HEIDI and were therefore not supported by multiple converging lines of evidence. Tier 2 classification reflects incomplete genetic triangulation rather than a lack of biological relevance, and these proteins remain of potential interest for future functional investigation.

### Network-based functional profiling

The global tissue-integrated network revealed a modest yet biologically informative interaction landscape among the genes encoding the 12 prioritised proteins (**Figure 3**). *STAT3* emerged as a central node, exhibiting extensive connectivity with multiple genes, including *STX4*, *RALB*, *SGSH*, and *NFKB1*, through physical interactions [2]. Upon incorporating approved pharmacological targets for psoriasis, *STAT3* and *NFKB1* emerged as hub nodes in the network, underscoring their pivotal role in orchestrating inflammatory signalling cascades relevant to disease pathogenesis. Both genes demonstrated strong interactions with the glucocorticoid receptor *NR3C1*. Moreover, the *SGSH,* a lysosomal sulfatase, exhibited notable co-expression patterns with several psoriasis drug targets in inflammatory tissues, such as *TYK2*, *IL17RA* and *PDE4A*. Its further interaction with *VDR* (vitamin D receptor), supporting the role of the latter as an immunometabolic mediator bridging hormone signalling with cytokine-driven inflammatory cascade. Peripheral nodes in the network included cytokine-related genes such as *FLT3*, *TNFSF11*, and *STX4,* all retained due to their established or putative roles in immune modulation. In contrast, *IL12B*, *IL18*, and *IL23A* were positioned at the margins of the network, indicative of more specialised or context-dependent immunoregulatory functions. A comprehensive summary of the enrichment results is provided in **Table S5**. Gene Ontology (GO) over-representation analysis demonstrated enrichment of cytokine-related biological processes among the Tier 1 proteins (**Figure S33**). In particular, pathways involving TNF superfamily cytokine production and regulation of immune cytokine signalling were significantly enriched after false discovery rate (FDR) correction. Proteins such as NFKB1, SPHK2, IL18, and STAT3 mapped to these modules, illustrating convergence of prioritised targets within established inflammatory signalling pathways relevant to psoriasis.

**Figure 3.**
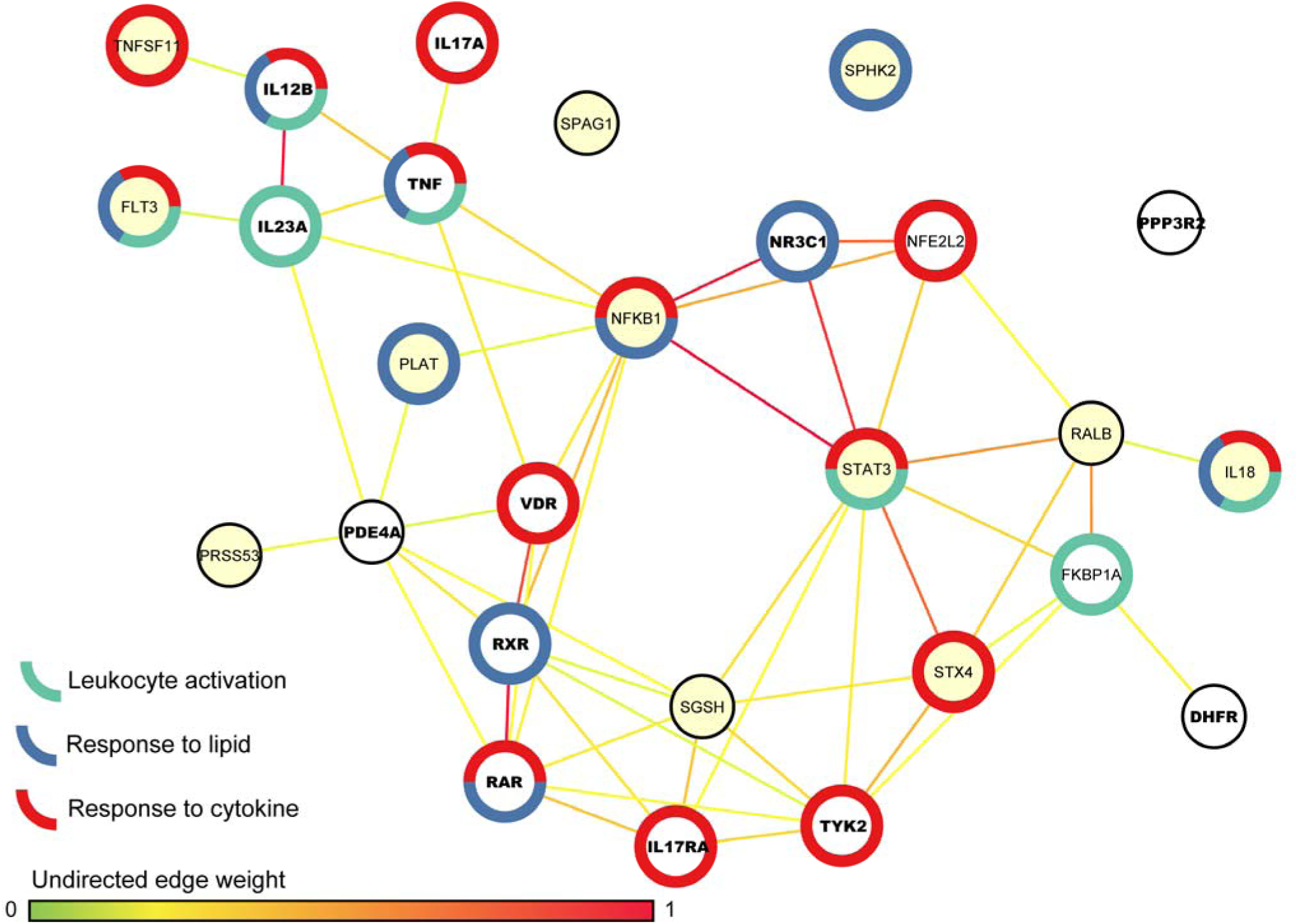
Protein-protein interactions (PPI) between prioritised genes. Each node corresponds to genes encoding the prioritised proteins. Nodes colored in yellow represent prioritised genes, while nodes coloured in white represent known drug targets for psoriasis. A discrete border colour scale was applied in nodes participating in selected biological processes. Nodes with bold font represent approved psoriasis drug targets.

### Druggability enrichment analysis

Genome for REPositioning drugs (GREP) enrichment analysis showed that genes encoding psoriasis-associated prioritised proteins were enriched in the targets of approved medications for ‘antineoplastic and immunomodulating agents’ in Anatomical Therapeutic Chemical (ATC) group L (OR=3.6, P_Fisher’s exact_=0.009) and ‘blood and blood forming organs’ in ATC group B (OR=4.5, P_Fisher’s exact_=0.038) (**Table 2**). Notably, this included *FLT3* (sunitinib, sorafenib, ponatinib, midostaurin), *NFKB1* (thalidomide, triflusal), *TNFSF11* (lenalidomide), and *PLAT* (urokinase, aminocaproic acid, conestat alfa).

**Table 2.**
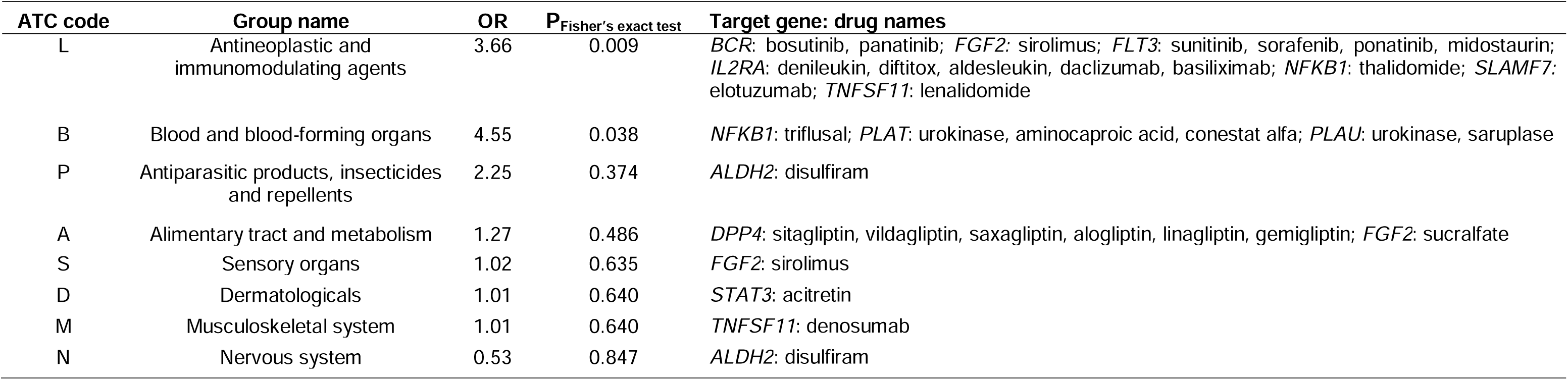
Enrichment of genes encoding proteins associated with psoriasis in the *cis*-MR analysis (P_2SMR_-_FDR_<0.05) in genes encoding ATC drug targets.

### Single-cell RNA-seq analysis of prioritised targets in psoriasis and IL-23 blockade

We evaluated the cell-type specificity of putative causal genes encoding prioritised proteins in healthy, psoriatic lesional, and non-lesional skin using single-cell RNA-seq datasets. Several genes (*XCL1*, *XCL2*, *IGLC2*, *TNFSF11*, *A2ML1*) exhibited cell-type–restricted expression patterns, whereas others (*STAT3*, *JUND*, *STX4*) were broadly expressed across epithelial and immune compartments (**Figure S34**). *XCL1* and *XCL2* were enriched in innate lymphoid cells and NK cells. *IL10* expression was higher in non-lesional compared with lesional psoriatic skin. In contrast, *NFKB1* expression was enriched in dendritic cells, macrophages, and T-cell subsets in lesional skin.

We next assessed transcriptional changes following IL-23 blockade (risankizumab). Five genes (*SPAG1*, *PRSS53*, *PLAT*, *DDX58*, *A2ML1*) were significantly downregulated across keratinocyte compartments (**Figure 4, Table S6**). Conversely, six genes (*STX4*, *STAT3*, *MMP12*, *MAP4K5*, *IL18*, *FLT3*) were significantly upregulated across selected epithelial, stromal, and immune cell populations. Downregulated genes were primarily observed in keratinocyte subsets, whereas upregulated genes showed cell-type–specific induction in fibroblasts, endothelial cells, macrophages, monocytes, and keratinocytes. These results demonstrate condition- and time-resolved transcriptional changes following IL-23 inhibition.

**Figure 4.**
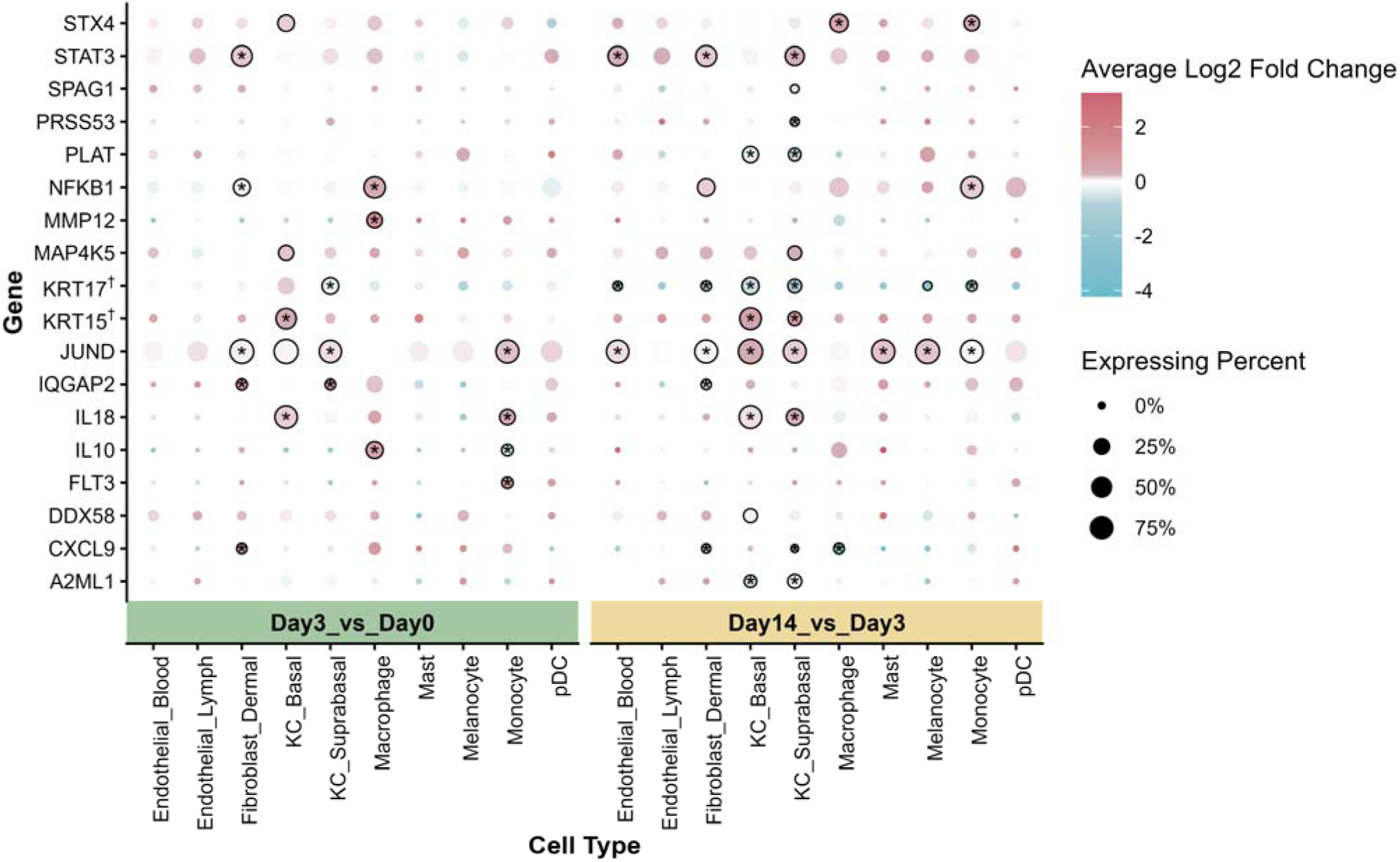
Cell-type–specific expression and treatment-associated transcriptional changes of prioritised genes. Differential expression of prioritised genes following IL-23 blockade (Day 3 vs Day 0 and Day 14 vs Day 3) across cell types. Colour scale denotes average log₂ fold change, and dot size indicates the percentage of expressing cells. Asterisks indicate statistically significant changes after multiple-testing correction. *KRT15* and *KRT17* genes are included as reference genes for comparison and are marked by †.

## DISCUSSION

This is a comprehensive cross-platform investigation of thousands of genetically predicted circulating plasma proteins in psoriasis, integrating large-scale proteomic datasets with the largest available psoriasis GWAS to prioritise putative causal targets. Through combined MR, colocalisation, and SMR-HEIDI analyses, twelve proteins (STX4, FLT3, NFKB1, IL18, PRSS53, SPAG1, SGSH, PLAT, RALB, TNFSF11, SPHK2, and STAT3) were prioritised as Tier 1 with convergent genetic support. Functional network and druggability analyses demonstrated enrichment of these targets within immune and inflammatory pathways and overlap with approved drug classes, particularly in ATC groups L and B. Complementary scRN-seq analyses further validated the cellular localisation and treatment-responsive modulation of several prioritised genes, linking genetic prioritisation with disease-relevant immune and epithelial compartments.

Our analyses prioritised STAT3 (signal transducer and activator of transcription 3), supported by several lines of evidence, including MR, colocalisation with gene expression in relevant tissues and differential expression in tissues from psoriasis patients compared to controls. STAT3 is a well-known transcription factor involved in cellular growth, differentiation, survival, and immune response (24). It has been implicated in diverse inflammatory and autoimmune conditions, including psoriasis (25), and several STAT3 inhibitors are already under investigation in preclinical and clinical trials. Preclinical studies have shown that inhibition of STAT3 signalling could lead to downregulation of pro-inflammatory cytokines (26,27) and improved skin pathology in psoriasis models. Consistent with these observations, our scRNA-seq data showed marked upregulation of *STAT3* in both lesional and non-lesional psoriatic keratinocytes, indicating broad activation across the epidermis. Moreover, mouse studies with keratinocyte-specific Stat3 deletion (K5-Stat3–/–) demonstrated reduced psoriasis-like dermatitis, while T cell–specific deletion had little effect, highlighting keratinocyte STAT3 as a key driver of psoriatic pathology (28).

Dendritic cells (DCs), as key antigen-presenting cells, play a central role in initiating and regulating immune responses and have been implicated in autoimmune diseases, including psoriasis (29). DCs process and present antigens to T cells and influence T-cell differentiation and effector responses (30). FMS-like tyrosine kinase 3 (FLT3) is a receptor tyrosine kinase expressed in hematopoietic progenitors that regulates DC development and lineage commitment, and its expression persists in mature DCs, supporting their immunological function (31,32). In psoriasis, FLT3⁺ dendritic cells accumulate in lesional skin, and pharmacological inhibition of FLT3 attenuates epidermal hyperplasia and inflammation in experimental models (33,34). Our genetic analyses extend this framework by providing MR and colocalisation evidence that elevated circulating FLT3 levels are associated with increased psoriasis risk. Consistently, our scRNA-seq analyses demonstrated robust upregulation of *FLT3* in DC1 populations in both non-lesional and lesional psoriatic skin, with the strongest induction observed in lesions. These findings support the relevance of FLT3⁺ dendritic cells in psoriasis pathobiology and suggest that enhanced myeloid signalling may contribute to disease amplification, although functional validation will be required to clarify the precise mechanisms involved.

Our study also highlighted evidence for PRSS53 (serine protease 53), a known psoriasis susceptibility locus (25). The protein is relatively understudied in the context of therapeutic targeting, but serine proteases are involved in skin barrier function, keratinocyte differentiation, and the regulation of inflammation. Dysregulation in these processes could potentially contribute to the pathogenesis of psoriasis (35). Similarly, STX4 (Syntaxin 4) has been previously identified as a psoriasis susceptibility loci in GWAS (36,37) and has been associated with mechanisms relevant to psoriasis such as the inflammatory response (38) and keratinocyte proliferation (39), but its specific role in psoriasis specific role in psoriasis remains largely unexplored. *SPAG1* represents a comparatively underexplored candidate in psoriasis. SPAG1 encodes a co-chaperone protein implicated in intracellular transport and cytoskeletal organisation. Although its role in inflammatory skin disease has not been well characterised, cytoskeletal dynamics and vesicular trafficking are central to keratinocyte differentiation and epidermal barrier maintenance (5,6). Given that aberrant keratinocyte proliferation and differentiation are hallmarks of psoriatic plaque formation (5), dysregulation of structural or trafficking pathways may contribute to disease pathogenesis. *SGSH* encodes N-sulfoglucosamine sulfohydrolase, a lysosomal enzyme involved in glycosaminoglycan degradation. Although primarily studied in lysosomal storage disorders, lysosomal function is increasingly recognised as an important regulator of immune-cell activation and inflammatory signalling. Lysosomal pathways intersect with antigen processing and cytokine regulation, processes highly relevant to psoriasis immunopathology (5,32). In our network analysis, *SGSH* demonstrated connectivity with established inflammatory mediators, suggesting a potential role in immune–metabolic regulation within psoriatic tissue.

Several other proteins demonstrated robust evidence of tissue-specific expression effects through SMR-HEIDI analysis and are noteworthy. NFKB1, which encodes the p50 subunit of the NF-κB transcription factor, is an established regulator of inflammatory and immune responses and has been implicated in multiple chronic inflammatory diseases (35). SGSH/CARD14 is another psoriasis susceptibility locus (25) and a potential activator of NF-κB signalling, a family of transcription factors regulating genes involved in immune and inflammatory responses (40). Evidence from mouse models supports a pathogenic role for NF-κB1 in psoriatic inflammation. In an imiquimod (IMQ)-induced psoriasis-like model, *Nfkb1*⁻/⁻ mice exhibited markedly attenuated skin inflammation, driven by reduced numbers and impaired proliferation of pro-inflammatory Vγ4⁺Vδ4⁺ γδT17 cells (41). IL18 (interleukin 18) is a pro-inflammatory cytokine regulating immune responses through induction of IFN-γ production in T cells and NK cells. Its expression is increased in psoriatic lesional skin compared to normal skin (42). Mouse IL-18 knockout models further support its pathogenic role: *Il-18* deficiency in IMQ-induced psoriasiform inflammation leads to reduced acanthosis and diminished dermal immune-cell infiltration, indicating that IL-18 promotes epidermal hyperplasia and inflammation (43). Together, these data highlight IL-18 as a driver of psoriatic inflammation.

*PLAT* (plasminogen activator, tissue type) encodes tissue-type plasminogen activator, a serine protease responsible for converting plasminogen to plasmin. Both plasminogen activators and plasmin have been detected in psoriatic lesions, and altered plasminogen–plasmin balance has been implicated in psoriasiform inflammation in murine models (44), suggesting a potential role for fibrinolytic pathways in epidermal hyperplasia and inflammatory amplification. *TNFSF11* encodes RANKL (Receptor Activator of Nuclear factor-kappa B Ligand), which is expressed in keratinocytes of inflamed skin (45) and is targeted by denosumab in osteoporosis. *SPHK2* (sphingosine kinase 2) has been implicated in immune regulation, including Th17 differentiation. In mouse models, pharmacological inhibition of SPHK2 attenuated psoriasis-like dermatitis, potentially through modulation of Th17 responses (46).

An important feature of our findings is the bidirectional pattern of association observed across prioritised proteins. Proteins whose genetically predicted higher levels were associated with higher psoriasis risk largely participate in immune activation and inflammatory amplification pathways, including dendritic cell development (FLT3) (29–32), NF-κB–mediated transcriptional regulation (NFKB1) (40,41), and cytokine-driven responses (IL18) (42). Conversely, proteins associated with reduced risk, such as RALB, TNFSF11, SPHK2, and STAT3, may reflect regulatory, compensatory, or context-dependent immunomodulatory processes that temper excessive inflammation or influence immune–keratinocyte crosstalk (45). SPHK2 has been implicated in Th17 modulation in psoriasis-like models (46), and STAT3 exerts pleiotropic, context-dependent effects across immune cell subsets (24,26). This bidirectional architecture is consistent with the tightly regulated yet dysbalanced immune network characteristic of psoriasis, in which pathogenic inflammatory drivers coexist with counter-regulatory mechanisms (5,7). Although MR provides evidence regarding the direction of genetically predicted effects, detailed mechanistic elucidation of these pathways requires further functional investigation.

A subset of the protein findings corresponds to previously reported psoriasis risk loci. For example, IL12B showed strong evidence of colocalisation with psoriasis (PP.H4=0.98), indicating a shared causal variant. TNFAIP3 and TNIP1 showed nominal MR associations (P<0.05) but did not pass the 5% FDR threshold and did not colocalise, with TNFAIP3 showing a high probability of distinct causal variants (PP.H3 = 0.99). IL23R and NFKBIA were not tested in MR due to insufficiently strong *cis*-pQTLs and showed no evidence of colocalization (PP.H3>PP.H4).

Our scRNA-seq analyses provide cellular context for the genetically prioritised targets and demonstrate that several of these genes are dynamically modulated during IL-23 blockade. The downregulation of *SPAG1*, *PRSS53*, *PLAT*, *DDX58*, and *A2ML1* across keratinocyte compartments is consistent with attenuation of inflammatory and stress-response programmes within the psoriatic epidermis. In particular, *DDX58* (RIG-I), an interferon-responsive sensor implicated in psoriatic IFN activation, and protease-associated genes such as *PLAT* and *A2ML1*, which contribute to tissue remodelling, showed coordinated reduction following treatment, suggesting partial normalisation of epidermal signalling. Conversely, the upregulation of *STX4*, *STAT3*, *MMP12*, *MAP4K5*, *IL18*, and *FLT3* across immune and stromal compartments indicates dynamic immune and tissue remodelling during therapeutic response. Increased *STAT3* expression in fibroblasts, endothelial cells, and suprabasal keratinocytes may reflect reparative or vascular remodelling processes, while induction of *MMP12* in macrophages may facilitate extracellular matrix turnover. Elevated *IL18* and *FLT3* expression in immune subsets may indicate reprogramming of inflammasome and myeloid signalling pathways as inflammation resolves.

The GREP-based enrichment analysis provides a hypothesis-generating perspective on potential drug repurposing. However, several of the enriched classes, particularly multikinase inhibitors targeting FLT3 and thalidomide analogues, are primarily developed for oncologic indications and have safety profiles that limit their suitability for long-term management of a chronic immune-mediated disease such as psoriasis (47–49). In addition, direct evaluation of these specific agents in established psoriasis animal models is limited. These findings are therefore best interpreted as reflecting convergence on shared immune and inflammatory pathways, including NF-κB– and cytokine-mediated signalling, rather than identifying immediate therapeutic candidates.

In conclusion, our study identified several circulating plasma proteins genetically associated with psoriasis, offering novel insights into disease pathogenesis and immune regulation. These findings highlight biologically relevant pathways that may inform future biomarker research and therapeutic investigation. However, further experimental and clinical validation is required to establish their biological relevance and translational potential in psoriasis.

### Strengths and limitations of the study

This study integrates cross-platform proteomic data with the largest psoriasis GWAS to date and applies a rigorous multi-layer causal inference framework, enhancing robustness and reducing bias. The inclusion of independent scRNA-seq analyses, including treatment-responsive datasets, further validates the biological and clinical relevance of prioritised targets. Nevertheless, several limitations should be acknowledged. First, although we restricted analyses to *cis*-pQTLs and applied sensitivity analyses, residual horizontal pleiotropy or unresolved linkage disequilibrium (LD) cannot be fully excluded. Second, interpretation of PP in colocalisation analysis requires caution; low PP.H4 values do not necessarily exclude shared causal variants, particularly in regions with limited statistical power or overlapping association signals. Third, the analysis was inherently limited to proteins with genome-wide significant pQTLs and available summary statistics, potentially omitting biologically relevant targets not captured by current proteomic platforms or genetic instruments. Fourth, only common variants (minor allele frequency ≥1%) were included, excluding rare variants that may exert stronger or context-specific effects. However, we successfully prioritised several proteins within the 16p11.2 locus, a genomic region previously implicated in psoriasis, supporting the validity of our approach. Fifth, some pQTLs, particularly those derived from the SomaScan platform, may be in high LD with protein-altering variants, raising the possibility of assay-related artefacts. Sixth, in loci such as the *PRSS53* region on chromosome 17, MR and colocalisation approaches may be insufficient to resolve the precise causal gene where multiple correlated candidates exist. Seventh, the absence of detailed clinical subtype information in the psoriasis GWAS meta-analysis precluded subtype-specific analyses. Finally, the genetic data used for the MR analyses were derived predominantly from individuals of European ancestry. Although this reduces heterogeneity due to population structure, residual population stratification may still bias MR estimates if genetic instruments correlate with ancestry-related factors influencing psoriasis risk, particularly within immune-related loci subject to selection pressures. While the contributing GWAS adjusted for principal components of ancestry, allele frequencies and linkage disequilibrium patterns may differ across populations, potentially affecting instrument validity and transferability.

## RESOURCE AVAILABILITY

### Lead contact

Further information and requests for resources should be directed to and will be fulfilled by the lead contact, Christos Chalitsios.

### Materials availability

Only publicly available data were utilised in this study. No new unique reagents or biological materials were generated.

### Data and code availability

The GWAS summary statistics for blood protein quantitative trait loci (pQTLs) from the UK Biobank Pharma Proteomics Project are available at http://ukb-ppp.gwas.eu/, and those from deCODE genetics can be accessed at https://www.decode.com/summarydata/. Summary statistics for psoriasis are publicly available via the GWAS Catalog under the study accession GCST90472771. Additional supporting information and data are available from the lead contact upon reasonable request.

## ACKNOWLEDGMENT

The authors would like to thank the staff and participants who contributed to the UK Biobank study and deCODE genetics, and the authors of the cited genome-wide association studies for sharing summary data. C.A. was financially supported by the “Andreas Mentzelopoulos Foundation”. This study was supported by the British Heart Foundation Research Excellence Award (RE/24/130023), UK Dementia Research Institute [UK DRI-5001], UK Medical Research Council, and NIHR Imperial Biomedical Research Centre.

## AUTHOR CONTRIBUTION

Conception and design: D.M, C.V.C, I.T

Development of methodology: D.M, C.V.C, A.D, I.T

Acquisition of data: D.M, C.V.C, J.H, N.M

Statistical analysis: D.M, C.V.C, J.H, A.S, N.M

Interpretation of data: All authors

Writing of the manuscript: D.M, C.V.C

Review and revision of the manuscript: All authors

Study supervision: I.T

D.M, C.V.C, J.H, N.M share first authorship.

## ETHICS STATEMENT

This study did not require ethical approval, as it was conducted using publicly available summary-level data from previously published genome-wide association studies (GWAS). All original studies obtained ethical approval from the relevant institutional review boards, and written informed consent was provided by all participants.

## STAR□METHODS

### KEY SOURCES TABLE

| REAGENT or RESOURCE | SOURCE | IDENTIFIER |
| --- | --- | --- |
| **Deposited data** |  |  |
| GWAS summary statistics for blood protein quantitative trait loci (pQTLs) from the UK Biobank Pharma Proteomics Project | Sun et al.<sup>16</sup> | http://ukb-ppp.gwas.eu/ |
| GWAS summary statistics for blood protein quantitative trait loci (pQTLs) from the deCODE genetics | Ferkingstad et al.<sup>15</sup> | https://www.decode.com/summarydata/ |
| GWAS summary statistics for psoriasis | Dand et al.<sup>50</sup> | [GCST90472771](https://www.gccr.org/gcst/GCST90472771) |
| Genotype-Tissue Expression Project (GTEx) v8 database | Li et al.<sup>62</sup> | https://gtexportal.org/home/ |
| GEO (GSE228421) | Francis et al.<sup>69</sup> | https://www.ncbi.nlm.nih.gov/geo/query/acc.cgi?acc=GSE228421 |
| **Software and algorithms** |  |  |
| R v4.3.2 |  | https://www.r-project.org/ |
| TwoSampleMR v0.5.6 |  | https://mrcieu.github.io/TwoSampleMR/ |
| Pair-Wise Conditional analysis and Colocalisation analysis | Robinson et al.<sup>59</sup> | https://github.com/jwr-git/pwcoco |
| Summary-data-based MR and HEIDI analyses | Zhu et al.<sup>61</sup> | https://yanglab.westlake.edu.cn/software/smr/#SMR&HEIDIanalysis |
| HumanBase platform | Greene et al.<sup>63</sup> | https://hb.flatironinstitute.org/ |
| Genome for REPositioning drugs version 1.0.0 | Sakaue et al.<sup>66</sup> | https://github.com/saorisakaue/GREP |

### DATA SOURSES

#### Proteomics and psoriasis GWAS

Plasma protein data were obtained from two large-scale proteomic GWAS. First, we utilised data from the UKB-PPP, where 2,940 protein targets (corresponding to 2,923 unique proteins) were quantified in 54,219 individuals of European ancestry using the Olink Explore 3072 platform, an antibody-based proximity extension assay (PEA) (16). Additionally, we included proteomic data from deCODE genetics, comprising 35,559 Icelandic participants. Using the SomaScan v4 platform, 4,907 aptamers targeting 4,719 unique proteins were measured (15). Summary statistics for psoriasis were obtained from the latest and largest genome-wide association meta-analysis of 18 GWAS containing 36,466 psoriasis cases and 458,078 controls of European ancestry (50).

## METHOD DETAILS

### Two-sample Mendelian randomisation

#### Genetic instruments of circulating plasma proteins

A two-sample MR analysis was conducted to evaluate the potential causal association between circulating protein levels and the risk of psoriasis. Genetic instruments were constructed using pQTLs identified in the aforementioned proteomic studies, derived separately from the UKB-PPP and the deCODE genetics datasets. We used *cis*-pQTLs, defined as single-nucleotide polymorphisms (SNPs) located within ±1 megabase (Mb) of the transcription start site of the gene encoding each protein (17). SNPs were included as instruments if they: (1) were associated with protein levels at genome-wide significance (p<5×10^-8^); (2) were located outside the extended major histocompatibility complex (MHC) region (chr6: 28.5-33.5 Mb (GRCh38) to avoid confounding due to complex LD; (3) were independent (r^2^<0.001) within a ±1 Mb window, based on LD pruning using a European ancestry reference panel from either 10,000 randomly selected UK Biobank participants or the 1000 Genomes Project (phase 3); and (4) had an F statistic≥10 (51), ensuring adequate instrument strength. SNPs meeting all criteria were retained for downstream MR analysis.

#### Quantification and statistical analysis

Estimates of the association between circulating plasma proteins and psoriasis were determined by the random-effects IVW method (52) when multiple instrumental SNPs were available, and the Wald ratio method when only a single SNP instrument was present. Sensitivity analyses were performed using the Cochran Q-test and the MR-Egger intercept test to assess the between-SNP association heterogeneity and horizontal pleiotropy (53,54). For proteins instrumented by more than two SNPs, MR results were considered robust if the weighted median and MR-Egger estimates were directionally consistent with the IVW estimate (55,56). Steiger filtering was applied to assess directionality, and SNPs explaining more variance in the outcome than in the exposure were excluded (57). Multiple testing was controlled using the Benjamini–Hochberg FDR at □ = 0·05. MR analysis was performed with R v4.3.2 statistical software.

### Colocalisation analysis

To address the possibility of horizontal pleiotropy due to genetic variants in LD (58), we performed pair-wise conditional analysis and colocalisation for protein–psoriasis pairs with prior evidence of association from MR (59). The analysis included common biallelic SNPs (minor allele frequency>0.01) located within a ±500 kb window around the transcription start site of the corresponding protein-coding gene. The pair-wise conditional analysis and colocalization model considers multiple causal variants within the same region and tests five mutually exclusive hypotheses: H0 (no association with either trait), H1 (association with the protein only), H2 (association with psoriasis only), H3 (distinct variants associated with each trait), and H4 (a shared variant associated with both traits). Default prior probabilities were used, assigning each SNP a probability of 1×10⁻□ of being causal for either trait and 1×10⁻□ of being a shared causal variant (60). A PP.H4>0.80 was considered strong evidence of colocalisation.

### Summary-data-based MR and HEIDI analyses

We conducted tissue-specific SMR analysis (61) for genes encoding the 27 proteins identified, focusing on the 27 genes using expression quantitative trait loci (eQTL) data from psoriasis-relevant tissues in the Genotype-Tissue Expression Project (GTEx) v8 database (62). Only genes with complete cis-eQTL summary statistics were included. To distinguish associations driven by a shared causal variant from those due to LD, we applied the heterogeneity in dependent instruments (HEIDI) test, which evaluates heterogeneity across multiple SNPs within a locus (63). The SMR and HEIDI analyses were performed using SMR software (SMR v1.3.1) (63). A P_SMR_<0.05 was defined as the significance level for SMR, and P_HEIDI_>0.05 was interpreted as evidence consistent with a shared causal variant rather than LD.

### Network-based functional profiling

To characterise functional modules associated with the prioritised proteins, the genome-scale integrated analysis of networks in tissues (GIANT) interface was employed through the HumanBase platform (64). GIANT aggregates 990 diverse genomic datasets, including direct interactions, co-expression networks and transcription factor binding sites, to enable a holistic interpretation of etiological mechanisms underlying complex traits. For this analysis, we applied the global network setting with a predefined interaction score threshold of ≥0.15 to ensure biologically meaningful connectivity. To enhance network interpretability and therapeutic relevance, we further expanded the interaction map by incorporating drug targets, as catalogued in DrugBank (65), from approved pharmacotherapies for psoriasis (7) (**Table S1**). Functional enrichment analysis was conducted using GO biological process terms via g: Profiler (66). Multiple testing was controlled using the Benjamini–Hochberg FDR at □ = 0·05.

### Druggability enrichment analysis

We utilised the GREP software version 1.0.0 (67) to perform druggability enrichment analyses on genes encoding psoriasis-associated proteins, aiming to identify drugs that target this gene set based on enrichment across clinical indication categories. As input, we included all proteins that surpassed a 5% FDR threshold in the two-sample MR analyses. Briefly, GREP performs Fisher’s exact test to determine whether the input gene set is significantly enriched for genes targeted by drugs, classified in clinical indication categories according to the ATC classification system. The software outputs a list of drugs associated with the input genes. GREP integrates curated data from two major drug-target databases: DrugBank (65) and the Therapeutic Target Database (68). A p-value < 0.05 (Fisher’s exact test) was considered statistically significant.

### Single-cell RNA-seq analysis of prioritised targets in psoriasis and IL-23 blockade

To provide independent biological validation of our prioritised targets, we analysed scRNA-seq data from healthy, non-lesional, and lesional psoriatic skin, and assessed transcriptional changes following IL-23 blockade (risankizumab) using longitudinal lesional scRNA-seq datasets. We utilised the processed AnnData object provided by Reynolds et al (69), which included author-derived cell-type annotations and harmonised metadata (E-MTAB-8142). For the treatment dataset (GSE228421), sample metadata were parsed to extract donor, biopsy site, and treatment timepoint (day 0, day 3, day 14) (70). Cells that failed the published QC thresholds and predicted doublets were removed, and the datasets were normalised and integrated using the SCTransform–RPCA workflow (70,71). Lineage identities were assigned by scoring canonical keratinocyte, fibroblast, myeloid/DC, T/NK, endothelial, and mast-cell marker sets from (70), and each cluster was labelled according to its highest-scoring lineage (71). For both datasets, a curated gene panel was intersected with the detected features, and cells were grouped using biologically relevant composite labels (cell type × condition or cell type × time point). For each gene–group pair, we calculated the percentage of cells expressing the gene (>0) and the raw count expression. Gene-wise min–max scaling was applied to produce matrices suitable for heatmaps and dot plots. Differential expression (DE) analysis was performed consistently across datasets (log-normalised expression) using Seurat’s Wilcoxon rank-sum test, with Benjamini–Hochberg FDR correction applied, and requiring a minimum of 3 cells per group (72). Comparisons included healthy versus lesional states (69) and Day 3 versus Day 0, Day 14 versus Day 3 (70), generating time- and condition-resolved transcriptional signatures.

