## Supplemental Figures for "Identifying protein biomarkers and therapeutic targets in psoriasis through integrative genomic, proteomic and transcriptomic analysis"

**Supplements**

**UK Biobank (Olink platform)**

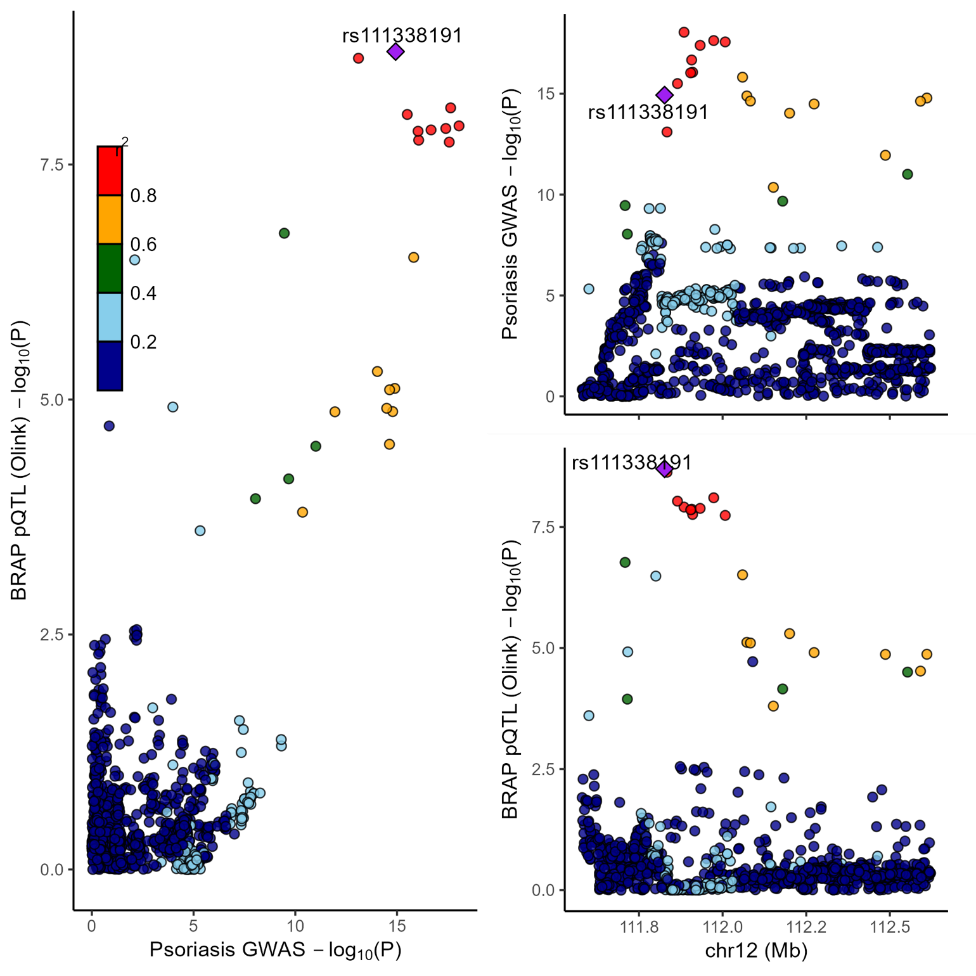

### **Figure S1.** Locus Compare plot. Left-hand panel represents psoriasis GWAS (y-axis) and blood pQTL for BRAP (x-axis) -log p values for the gene variants at the BRAP locus. Right-hand panels represent association data with the -log p values on the y-axis for the pQTL (BRAP) (upper panel) and psoriasis GWAS (lower panel) according to genomic coordinates (x-axis) spanning 1Mb centered on BRAP.

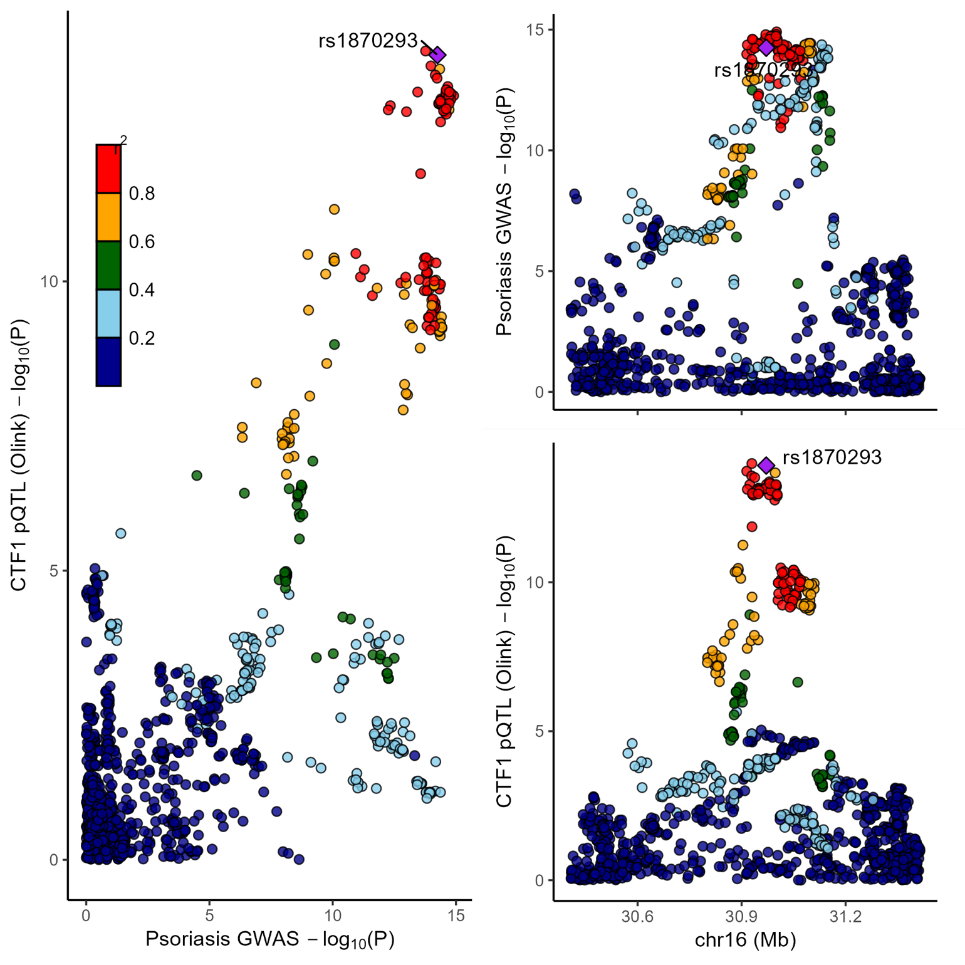

**Figure S2**. Locus Compare plot. Left-hand panel represents psoriasis GWAS (y-axis) and blood pQTL for CTF1 (x-axis) -log p values for the gene variants at the CTF1 locus. Right-hand panels represent association data with the -log p values on the y-axis for the pQTL (CTF1) (upper panel) and psoriasis GWAS (lower panel) according to genomic coordinates (x-axis) spanning 1Mb centered on CTF1.

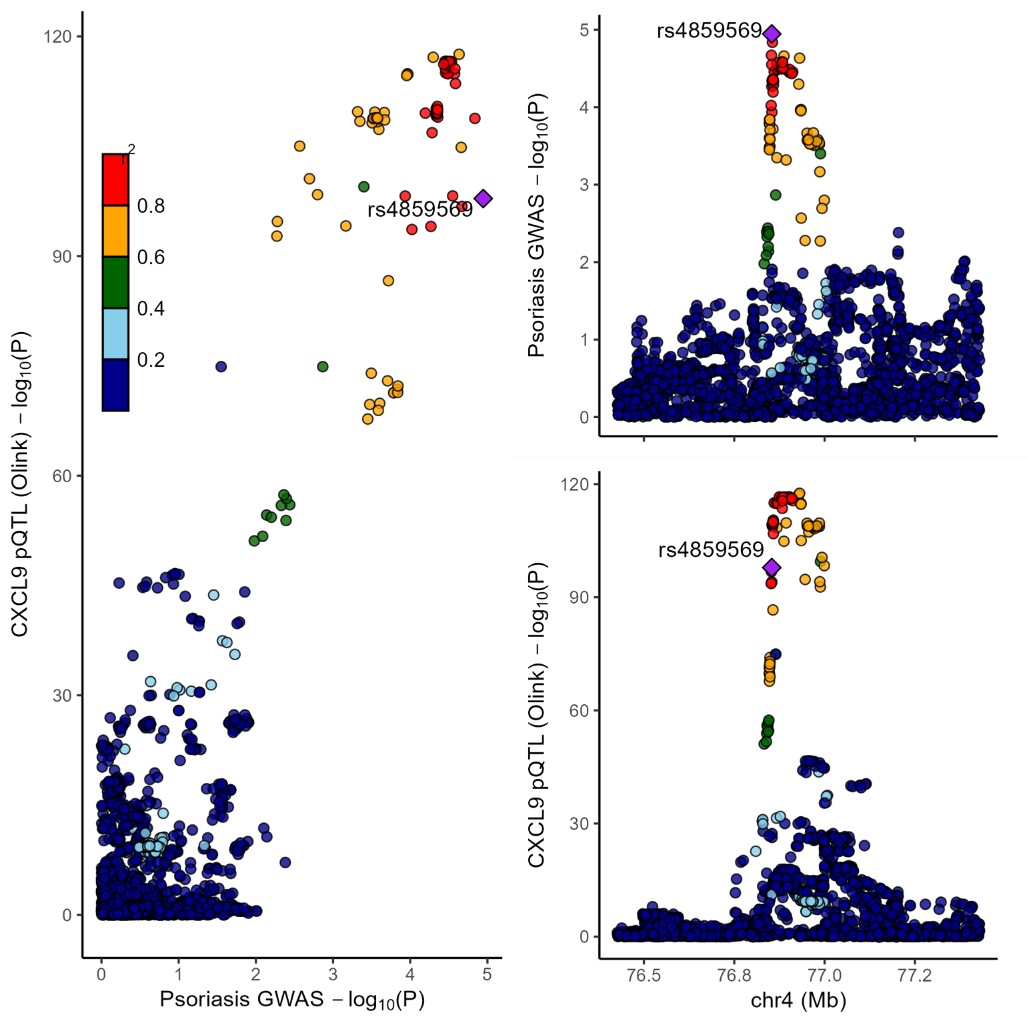

**Figure S3**. Locus Compare plot. Left-hand panel represents psoriasis GWAS (y-axis) and blood pQTL for CXCL9 (x-axis) -log p values for the gene variants at the CXCL9 locus. Right-hand panels represent association data with the -log p values on the y-axis for the pQTL (CXCL9) (upper panel) and psoriasis GWAS (lower panel) according to genomic coordinates (x-axis) spanning 1Mb centered on CXCL9.

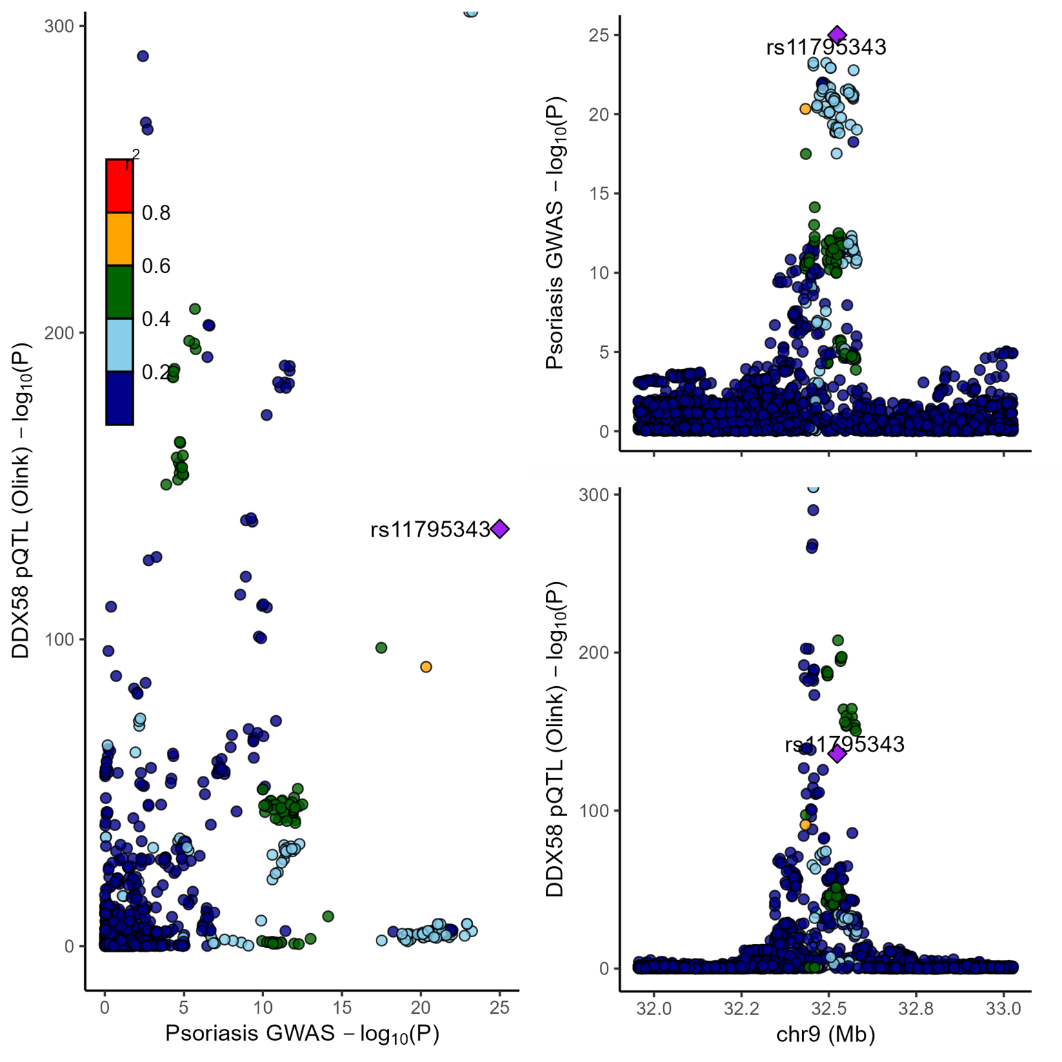

**Figure S4**. Locus Compare plot. Left-hand panel represents psoriasis GWAS (y-axis) and blood pQTL for DDX58 (x-axis) -log p values for the gene variants at the DDX58 locus. Right-hand panels represent association data with the -log p values on the y-axis for the pQTL (DDX58) (upper panel) and psoriasis GWAS (lower panel) according to genomic coordinates (x-axis) spanning 1Mb centered on DDX58.

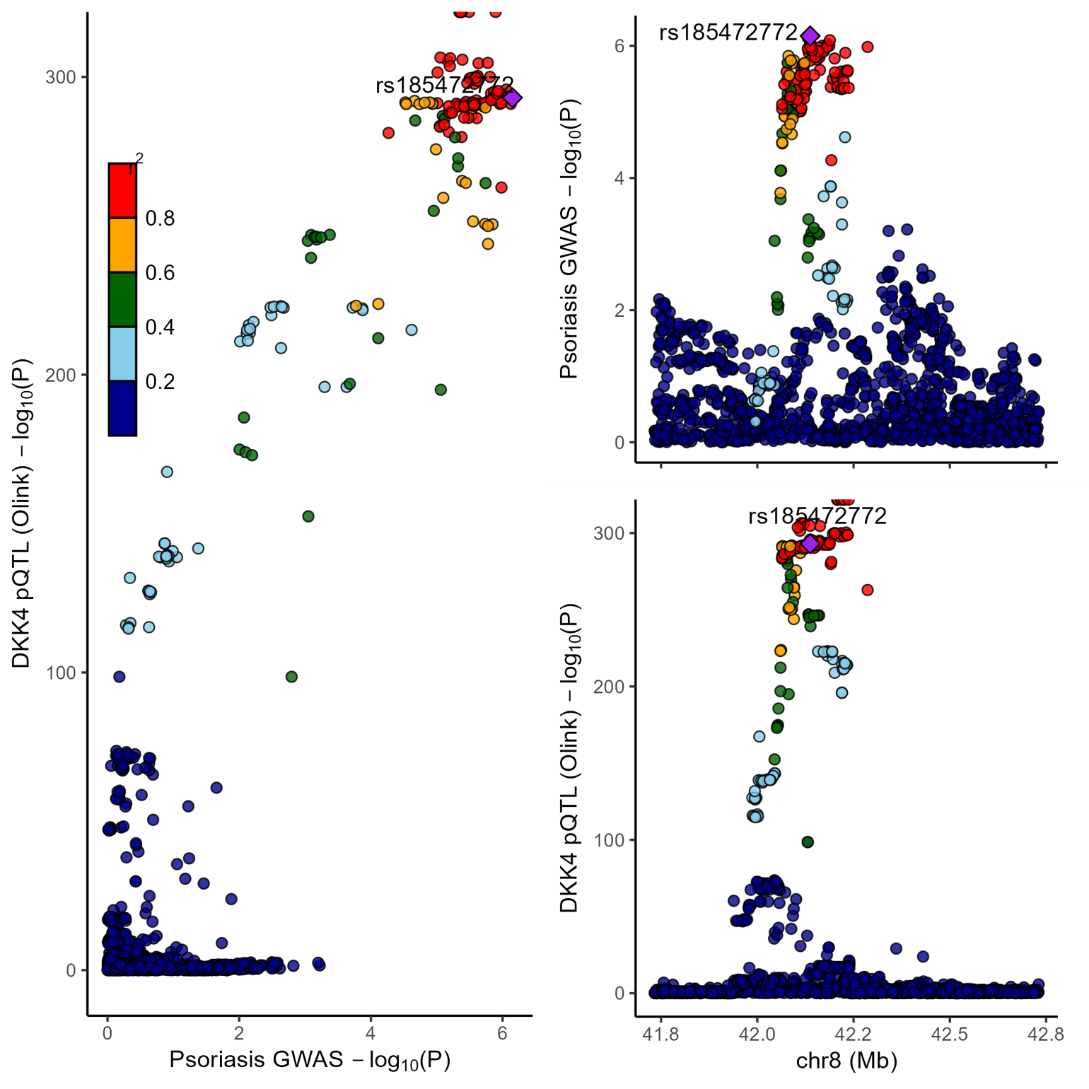

**Figure S5**. Locus Compare plot. Left-hand panel represents psoriasis GWAS (y-axis) and blood pQTL for DKK4 (x-axis) -log p values for the gene variants at the DKK4 locus. Right-hand panels represent association data with the -log p values on the y-axis for the pQTL (DKK4) (upper panel) and psoriasis GWAS (lower panel) according to genomic coordinates (x-axis) spanning 1Mb centered on DKK4.

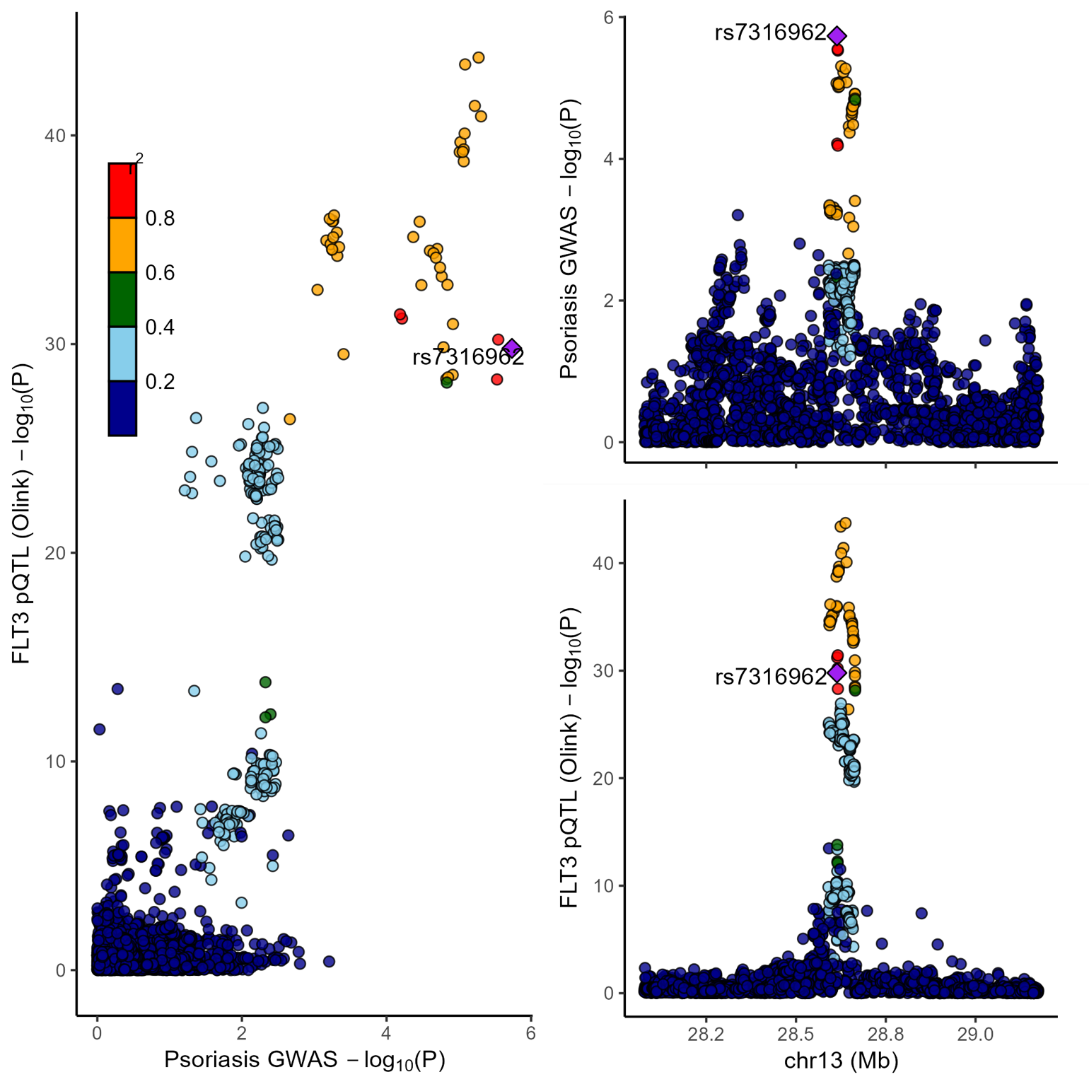

**Figure S6**. Locus Compare plot. Left-hand panel represents psoriasis GWAS (y-axis) and blood pQTL for FLT3 (x-axis) -log p values for the gene variants at the FLT3 locus. Right-hand panels represent association data with the -log p values on the y-axis for the pQTL (FLT3) (upper panel) and psoriasis GWAS (lower panel) according to genomic coordinates (x-axis) spanning 1Mb centered on FLT3.

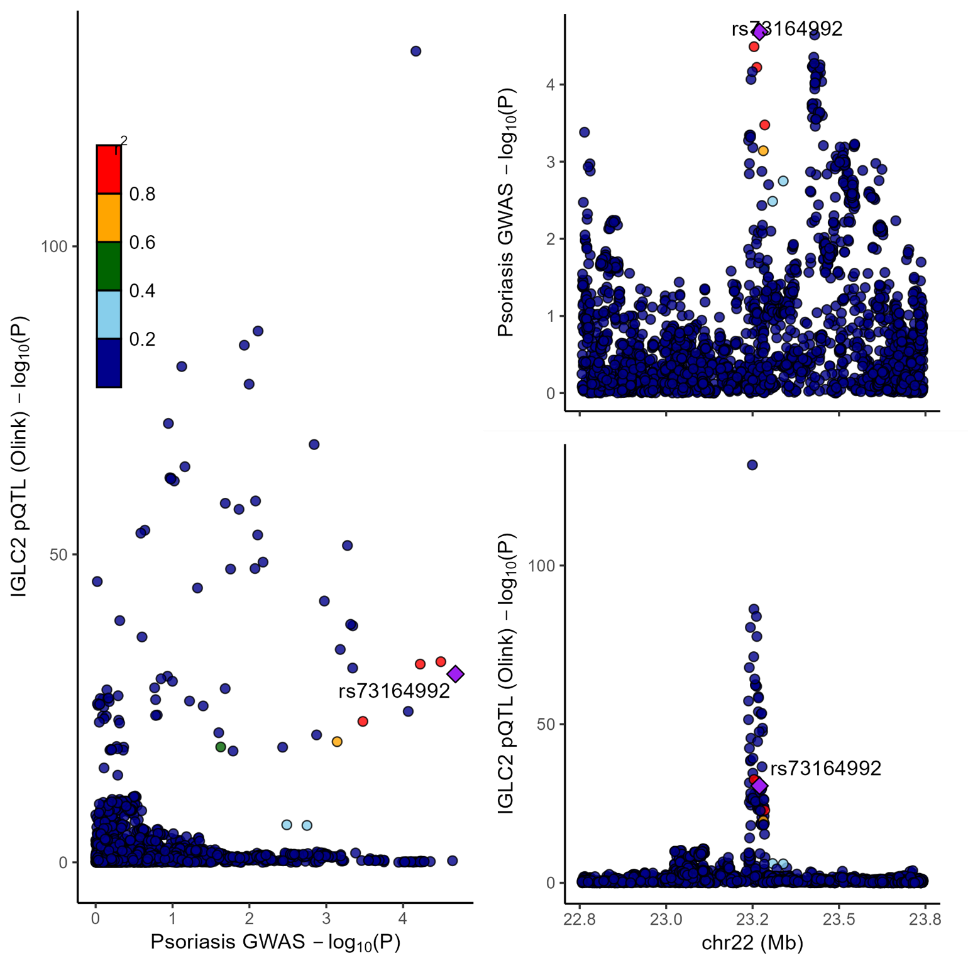

**Figure S7.** Locus Compare plot. Left-hand panel represents psoriasis GWAS (y-axis) and blood pQTL for IGLC2 (x-axis) -log p values for the gene variants at the IGLC2 locus. Right-hand panels represent association data with the -log p values on the y-axis for the pQTL (IGLC2) (upper panel) and psoriasis GWAS (lower panel) according to genomic coordinates (x-axis) spanning 1Mb centered on IGLC2.

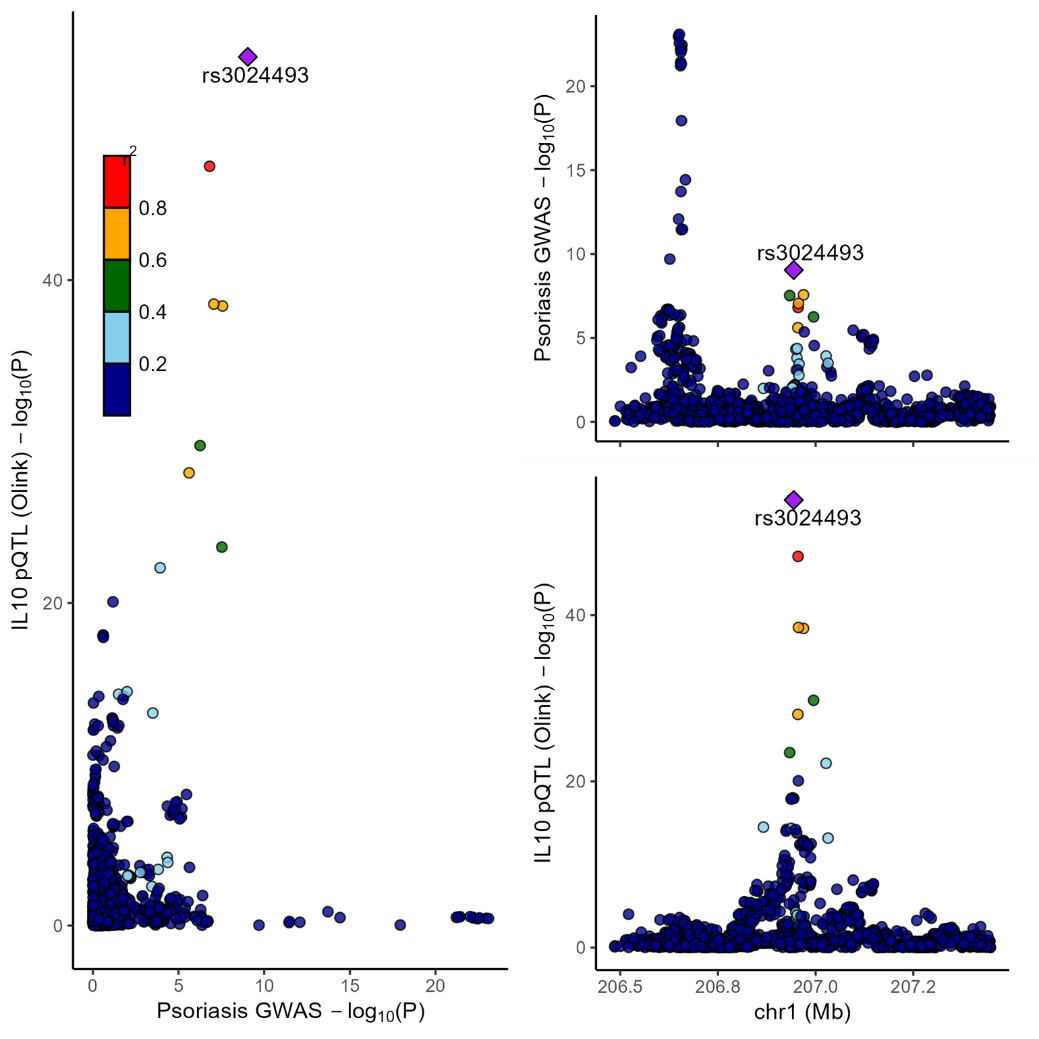

**Figure S8**. Locus Compare plot. Left-hand panel represents psoriasis GWAS (y-axis) and blood pQTL for IL10 (x-axis) -log p values for the gene variants at the IL10 locus. Right-hand panels represent association data with the -log p values on the y-axis for the pQTL (IL10) (upper panel) and psoriasis GWAS (lower panel) according to genomic coordinates (x-axis) spanning 1Mb centered on IL10.

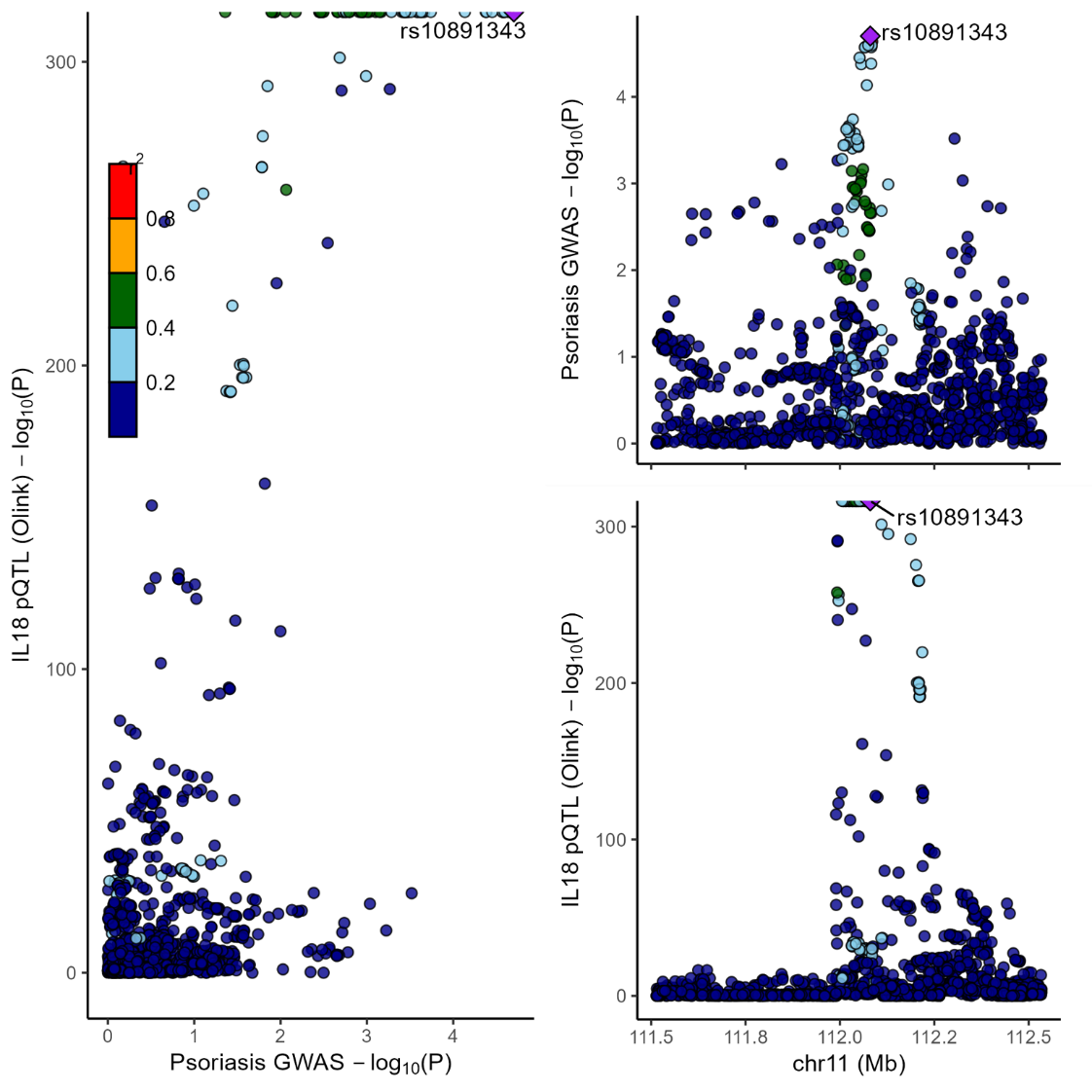

**Figure S9**. Locus Compare plot. Left-hand panel represents psoriasis GWAS (y-axis) and blood pQTL for IL18 (x-axis) -log p values for the gene variants at the IL18 locus. Right-hand panels represent association data with the -log p values on the y-axis for the pQTL (IL18) (upper panel) and psoriasis GWAS (lower panel) according to genomic coordinates (x-axis) spanning 1Mb centered on IL18.

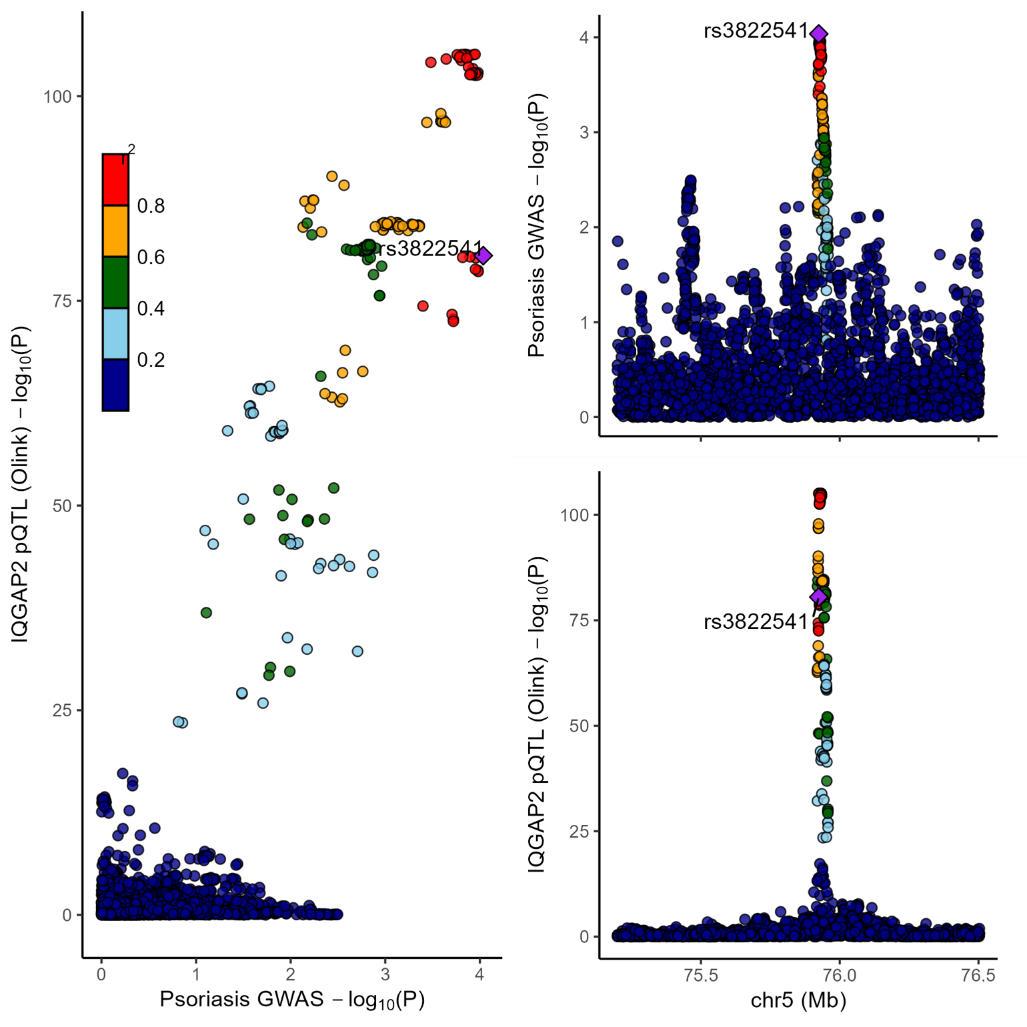

**Figure S10**. Locus Compare plot. Left-hand panel represents psoriasis GWAS (y-axis) and blood pQTL for IQGAP2 (x-axis) -log p values for the gene variants at the IQGAP2 locus. Right-hand panels represent association data with the -log p values on the y-axis for the pQTL (IQGAP2) (upper panel) and psoriasis GWAS (lower panel) according to genomic coordinates (x-axis) spanning 1Mb centered on IQGAP2.

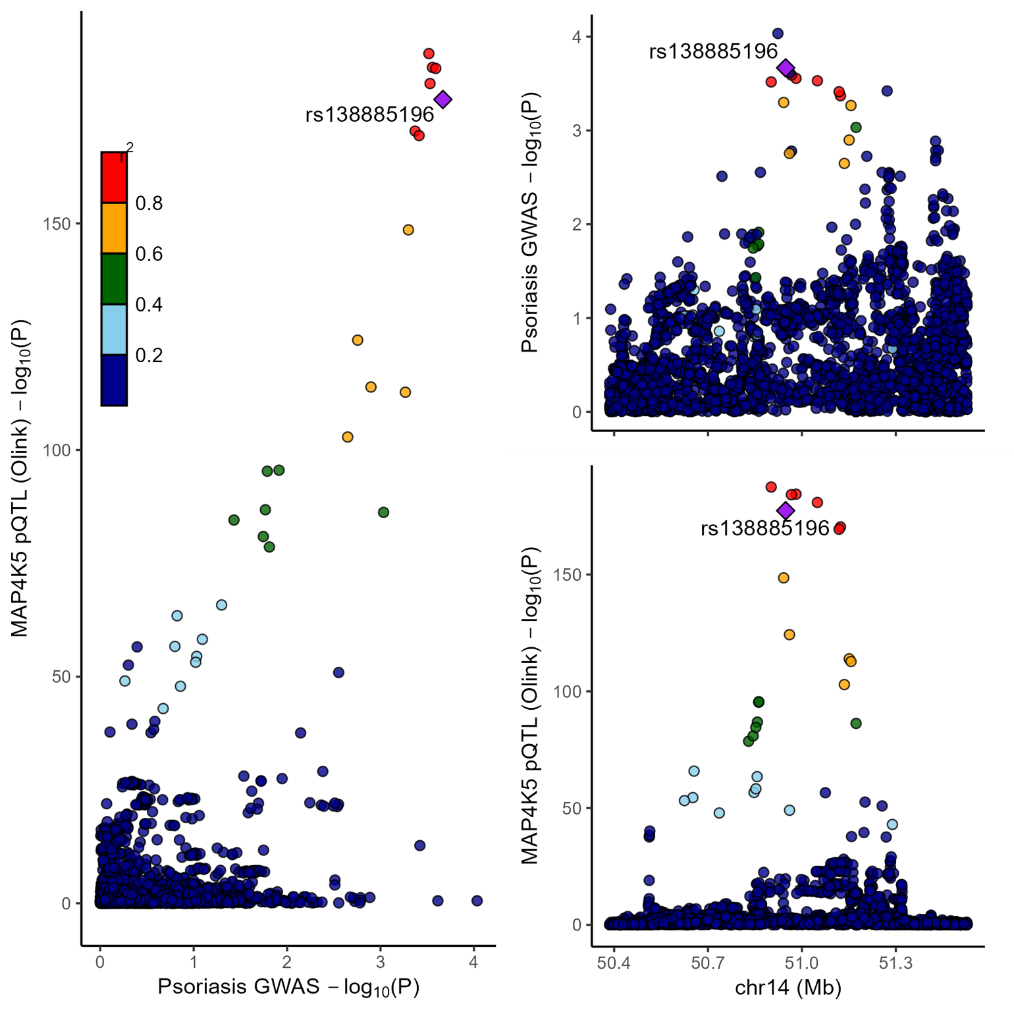

**Figure S11**. Locus Compare plot. Left-hand panel represents psoriasis GWAS (y-axis) and blood pQTL for MAP4K5 (x-axis) -log p values for the gene variants at the MAP4K5 locus. Right-hand panels represent association data with the -log p values on the y-axis for the pQTL (MAP4K5) (upper panel) and psoriasis GWAS (lower panel) according to genomic coordinates (x-axis) spanning 1Mb centered on MAP4K5.

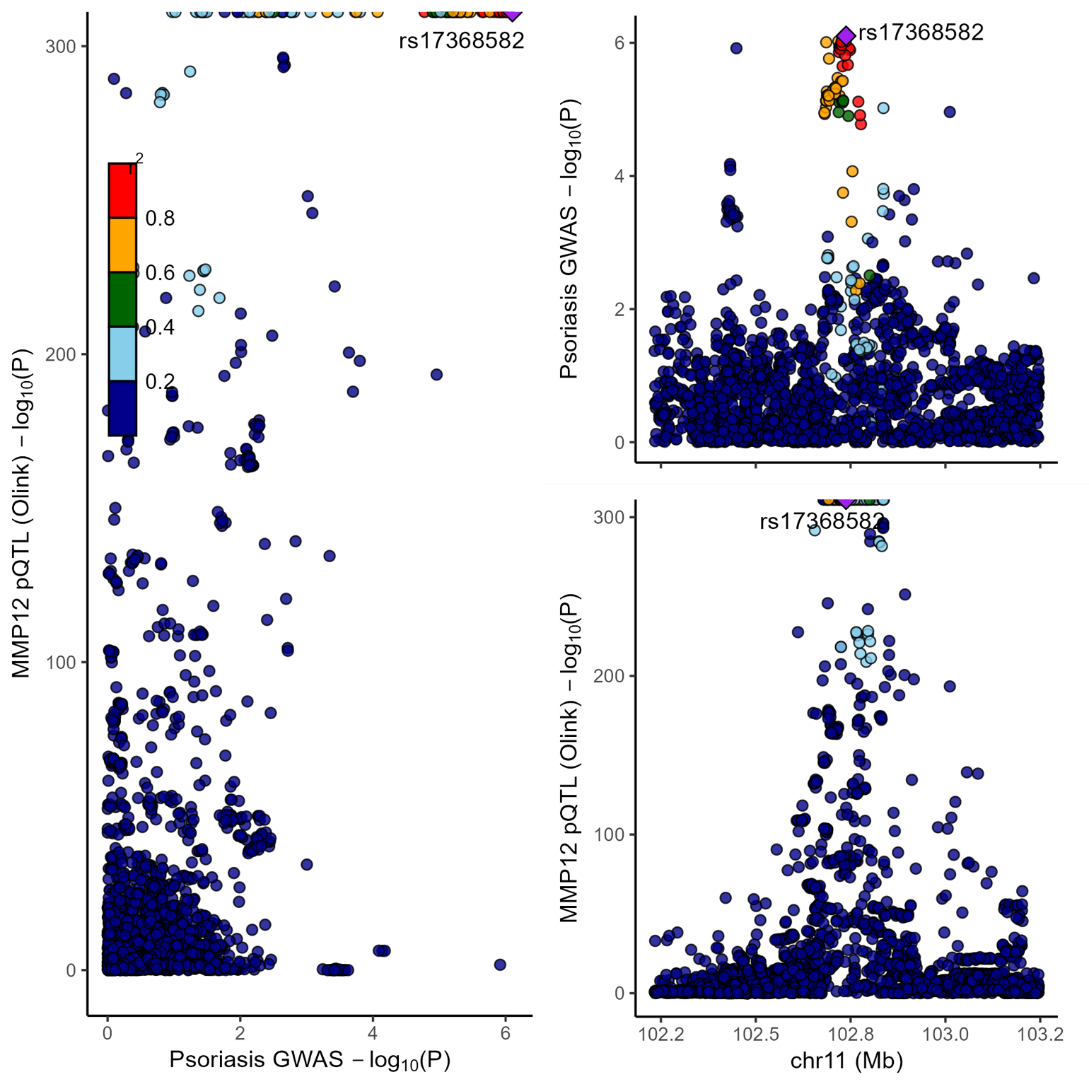

**Figure S12.** Locus Compare plot. Left-hand panel represents psoriasis GWAS (y-axis) and blood pQTL for MMP12 (x-axis) -log p values for the gene variants at the MMP12 locus. Right-hand panels represent association data with the -log p values on the y-axis for the pQTL (MMP12) (upper panel) and psoriasis GWAS (lower panel) according to genomic coordinates (x-axis) spanning 1Mb centered on MMP12.

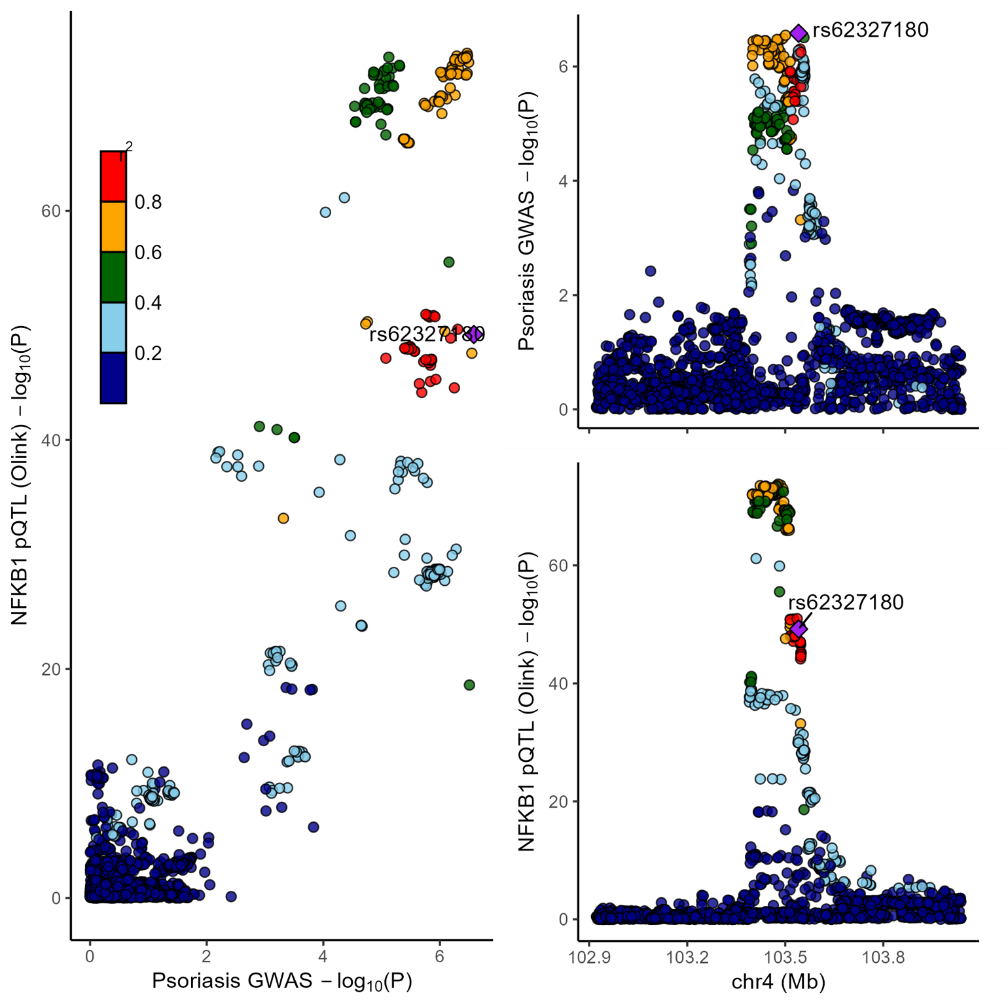

**Figure S13.** Locus Compare plot. Left-hand panel represents psoriasis GWAS (y-axis) and blood pQTL for NFKB1 (x-axis) -log p values for the gene variants at the NFKB1 locus. Right-hand panels represent association data with the -log p values on the y-axis for the pQTL (NFKB1) (upper panel) and psoriasis GWAS (lower panel) according to genomic coordinates (x-axis) spanning 1Mb centered on NFKB1.

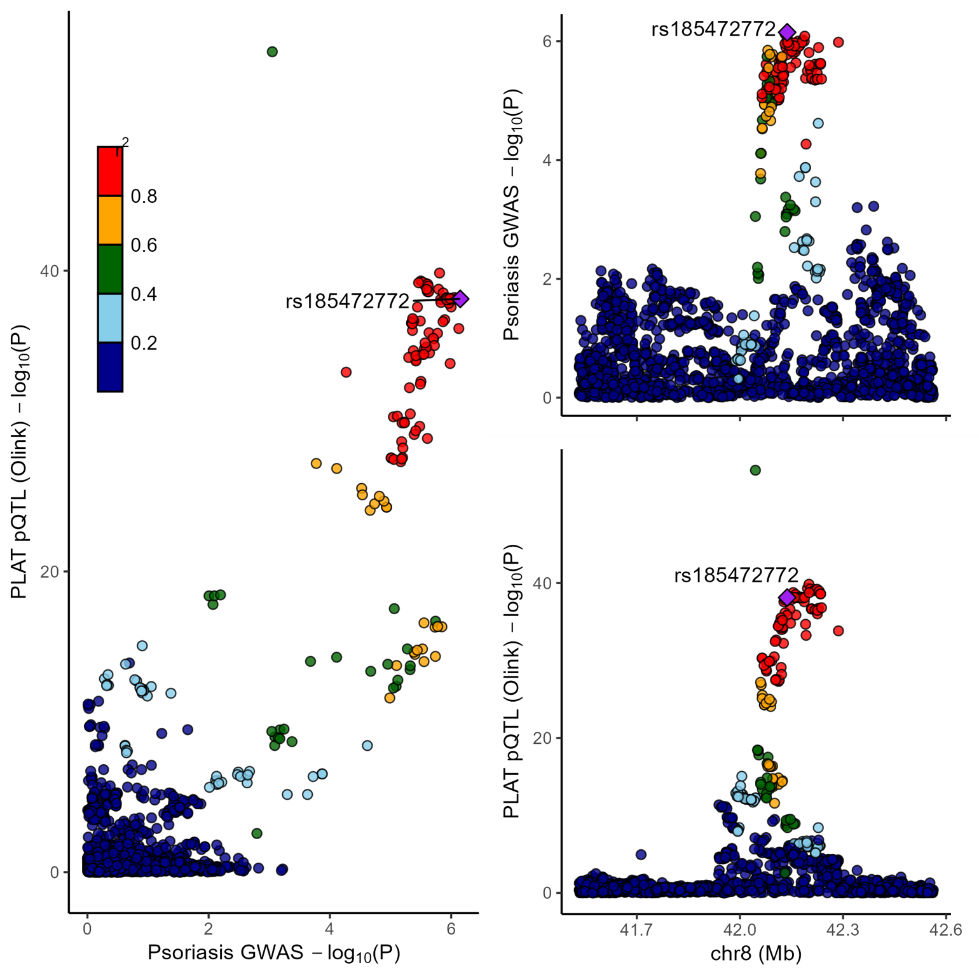

**Figure S14.** Locus Compare plot. Left-hand panel represents psoriasis GWAS (y-axis) and blood pQTL for PLAT (x-axis) -log p values for the gene variants at the PLAT locus. Right-hand panels represent association data with the -log p values on the y-axis for the pQTL (PLAT) (upper panel) and psoriasis GWAS (lower panel) according to genomic coordinates (x-axis) spanning 1Mb centered on PLAT.

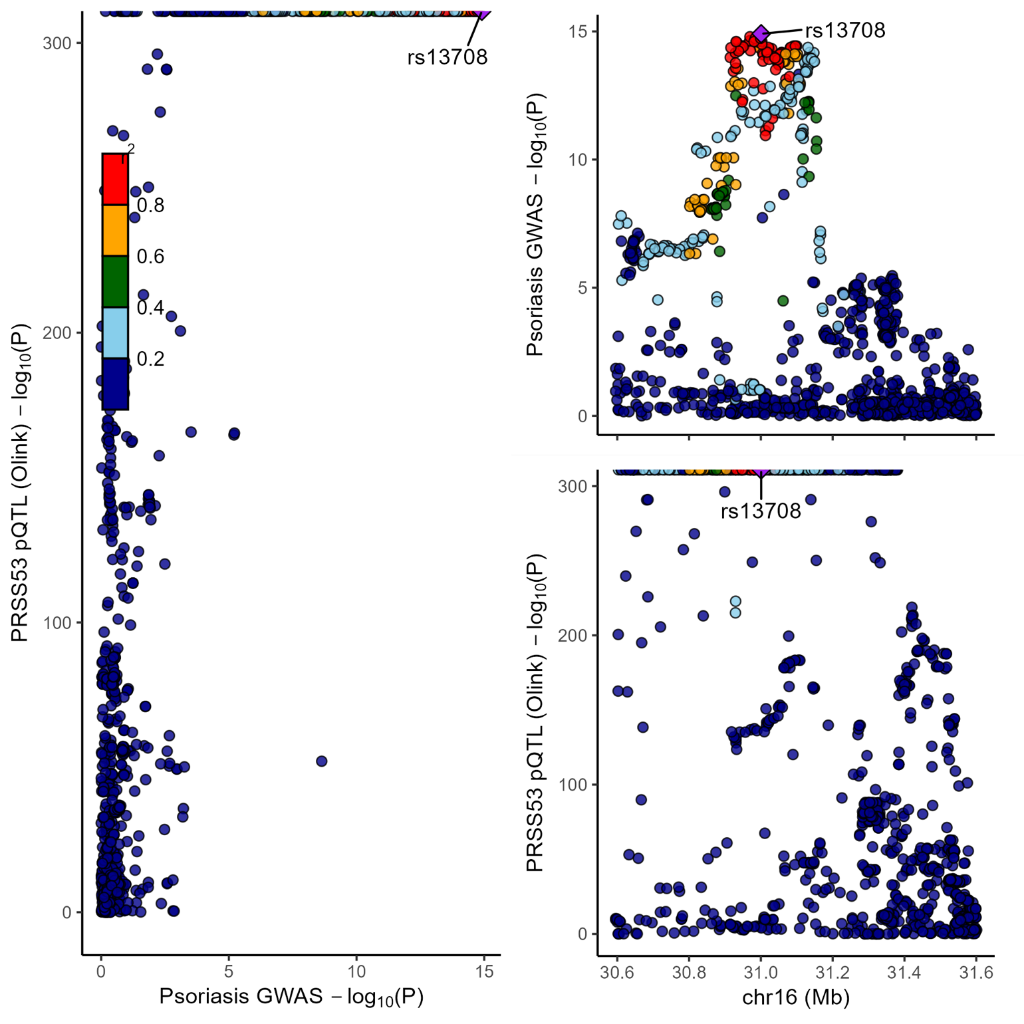

**Figure S15**. Locus Compare plot. Left-hand panel represents psoriasis GWAS (y-axis) and blood pQTL for PRSS53 (x-axis) -log p values for the gene variants at the PRSS53 locus. Right-hand panels represent association data with the -log p values on the y-axis for the pQTL (PRSS53) (upper panel) and psoriasis GWAS (lower panel) according to genomic coordinates (x-axis) spanning 1Mb centered on PRSS53.

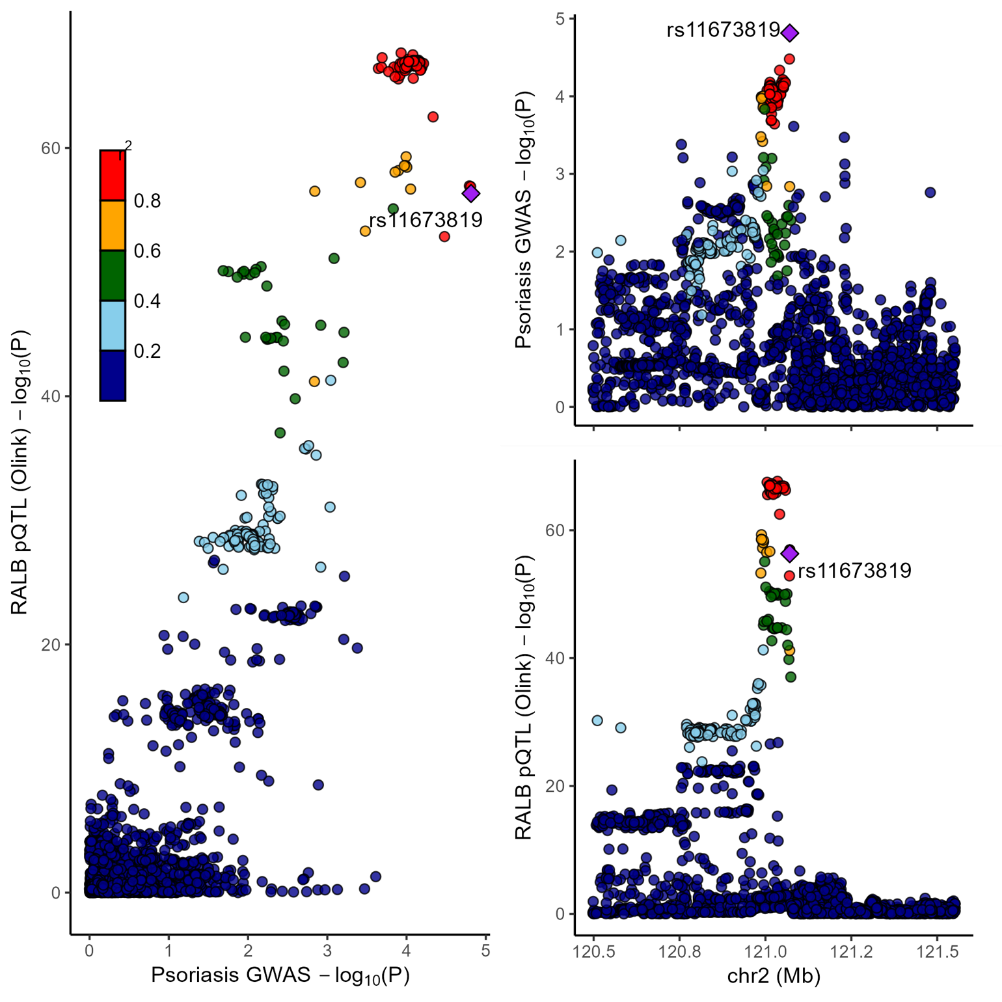

**Figure S16**. Locus Compare plot. Left-hand panel represents psoriasis GWAS (y-axis) and blood pQTL for RALB (x-axis) -log p values for the gene variants at the RALB locus. Right-hand panels represent association data with the -log p values on the y-axis for the pQTL (RALB) (upper panel) and psoriasis GWAS (lower panel) according to genomic coordinates (x-axis) spanning 1Mb centered on RALB.

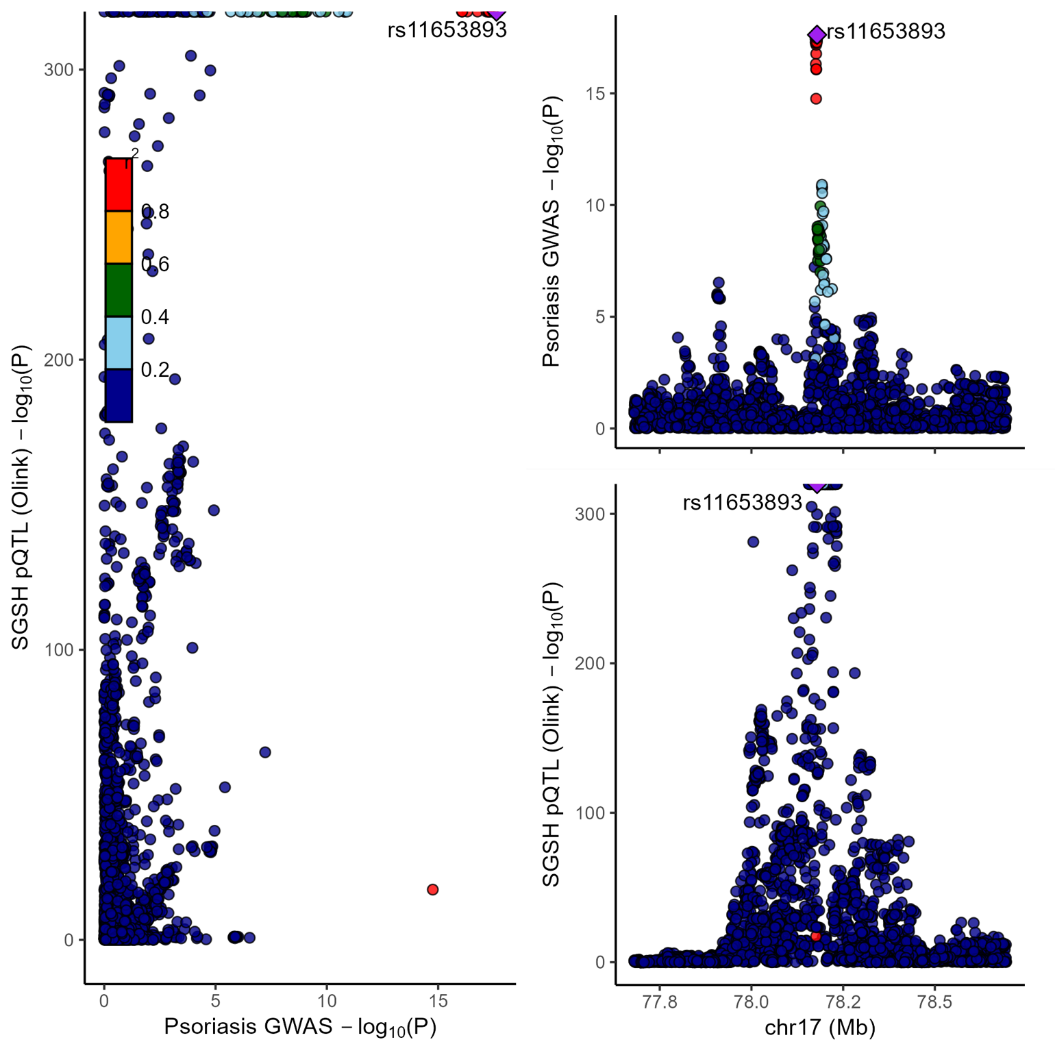

**Figure S17**. Locus Compare plot. Left-hand panel represents psoriasis GWAS (y-axis) and blood pQTL for SGSH (x-axis) -log p values for the gene variants at the SGSH locus. Right-hand panels represent association data with the -log p values on the y-axis for the pQTL (SGSH) (upper panel) and psoriasis GWAS (lower panel) according to genomic coordinates (x-axis) spanning 1Mb centered on SGSH.

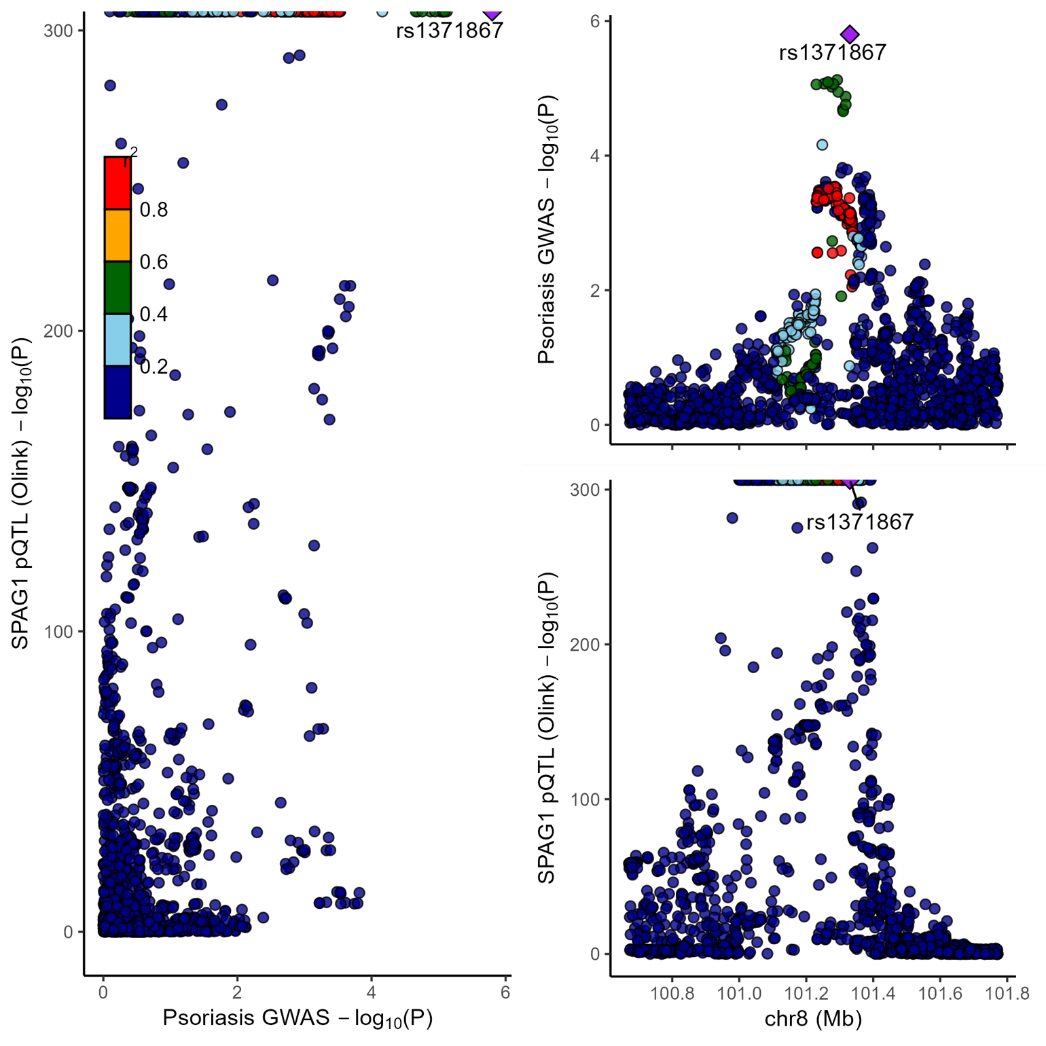

**Figure S18.** Locus Compare plot. Left-hand panel represents psoriasis GWAS (y-axis) and blood pQTL for SPAG1 (x-axis) -log p values for the gene variants at the SPAG1 locus. Right-hand panels represent association data with the -log p values on the y-axis for the pQTL (SPAG1) (upper panel) and psoriasis GWAS (lower panel) according to genomic coordinates (x-axis) spanning 1Mb centered on SPAG1.

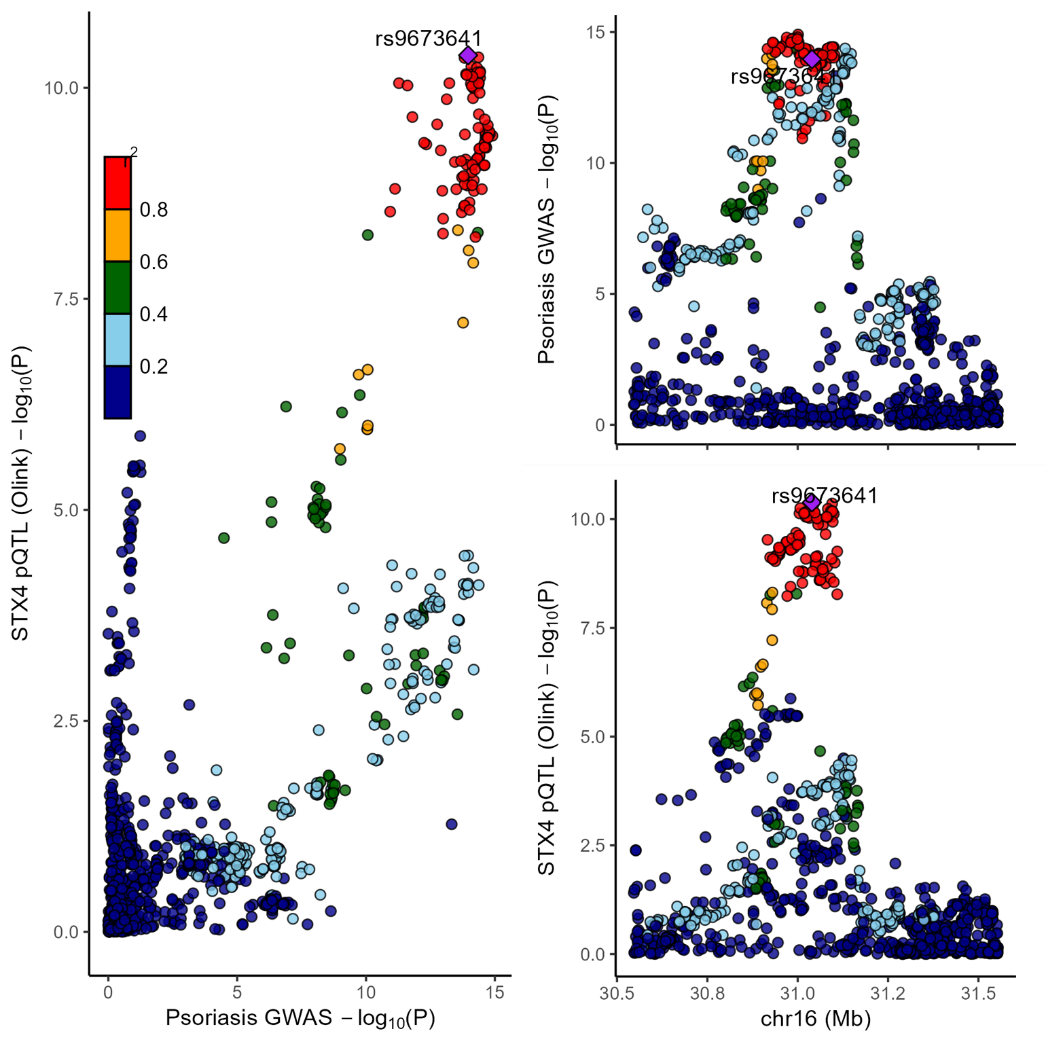

**Figure S19**. Locus Compare plot. Left-hand panel represents psoriasis GWAS (y-axis) and blood pQTL for STX4 (x-axis) -log p values for the gene variants at the STX4 locus. Right-hand panels represent association data with the -log p values on the y-axis for the pQTL (STX4) (upper panel) and psoriasis GWAS (lower panel) according to genomic coordinates (x-axis) spanning 1Mb centered on STX4.

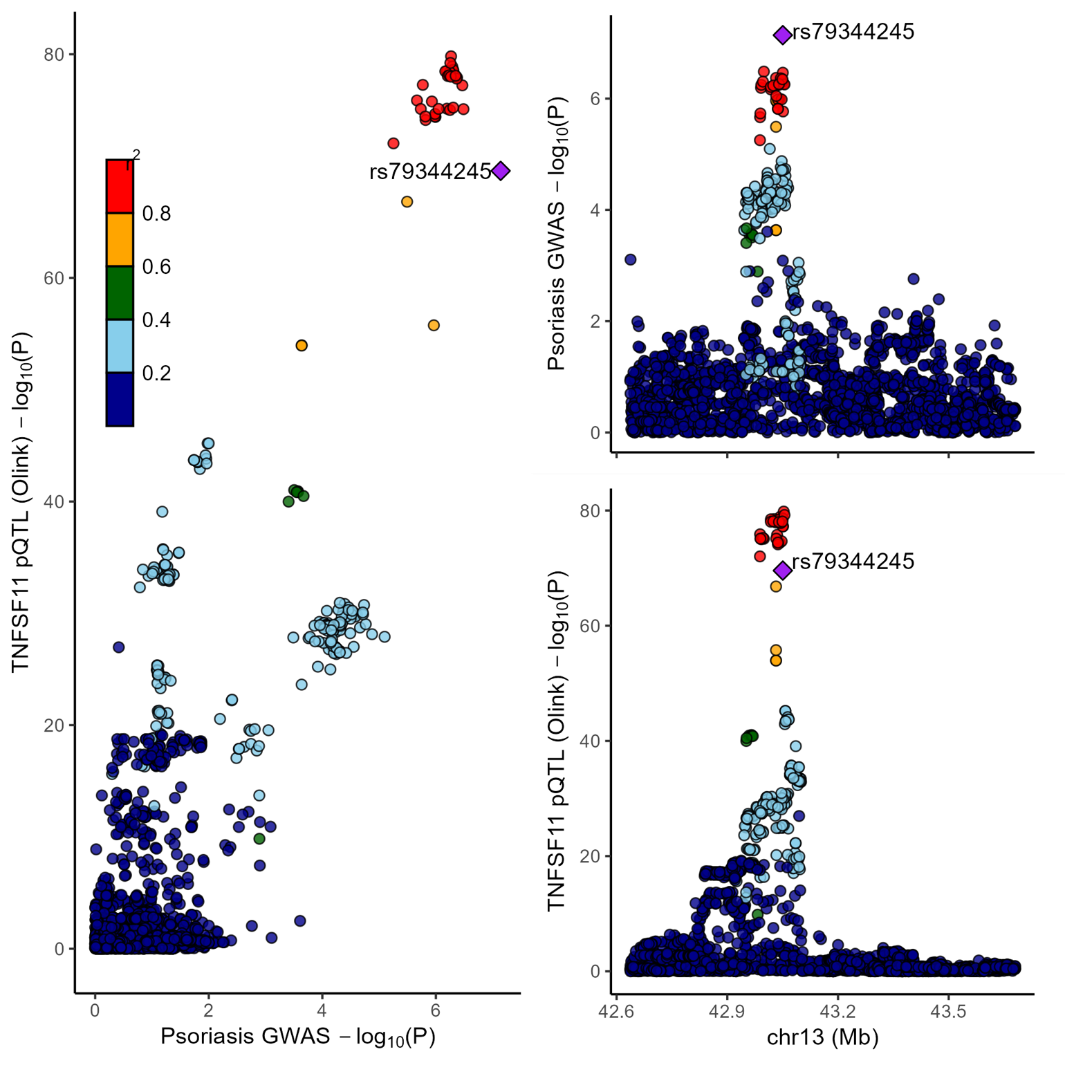

Figure 20.

**Figure S21**. Locus Compare plot. Left-hand panel represents psoriasis GWAS (y-axis) and blood pQTL for TNFSF11 (x-axis) -log p values for the gene variants at the TNFSF11 locus. Right-hand panels represent association data with the -log p values on the y-axis for the pQTL (TNFSF11) (upper panel) and psoriasis GWAS (lower panel) according to genomic coordinates (x-axis) spanning 1Mb centered on TNFSF11.

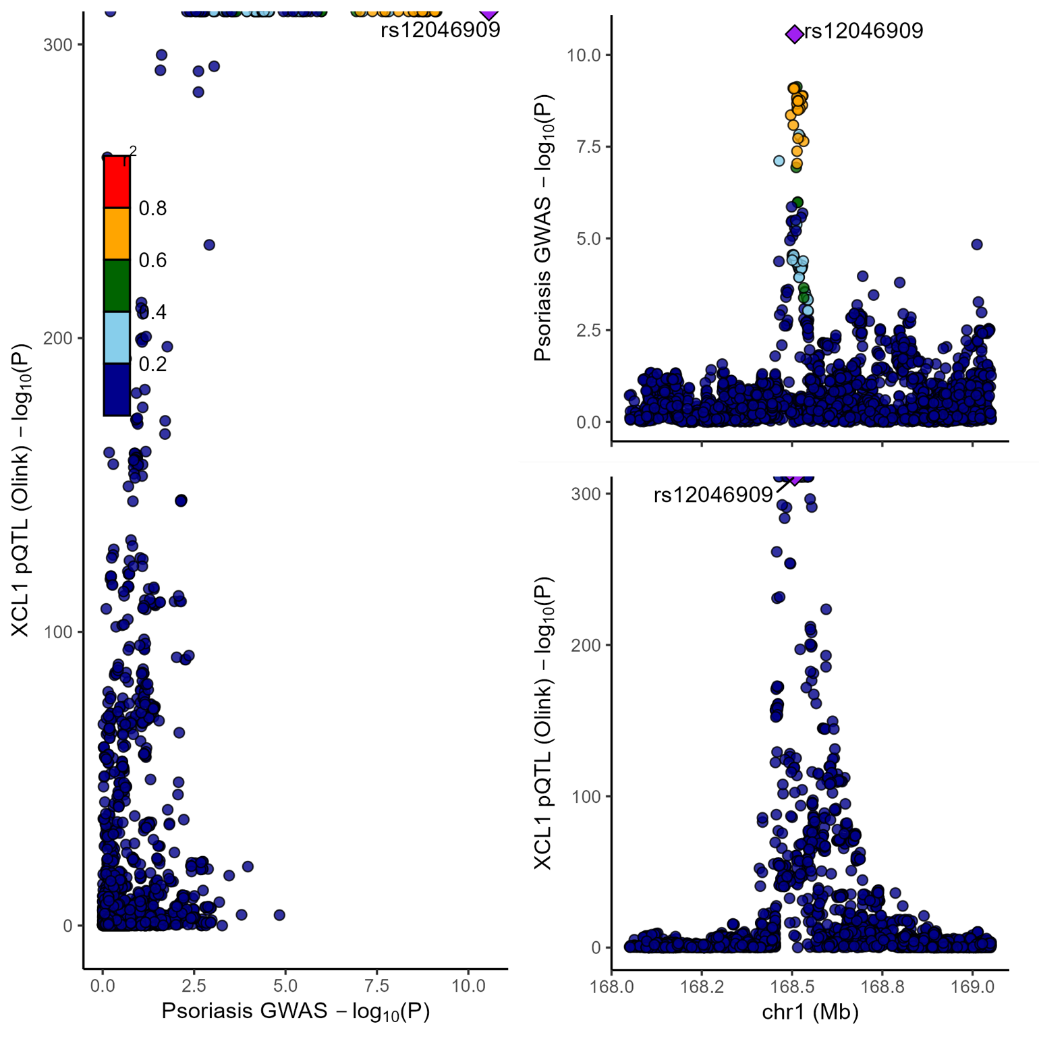

**Figure S22**. Locus Compare plot. Left-hand panel represents psoriasis GWAS (y-axis) and blood pQTL for XCL1 (x-axis) -log p values for the gene variants at the XCL1 locus. Right-hand panels represent association data with the -log p values on the y-axis for the pQTL (XCL1) (upper panel) and psoriasis GWAS (lower panel) according to genomic coordinates (x-axis) spanning 1Mb centered on XCL1.

**deCODE genetics (SomaScan platform)**

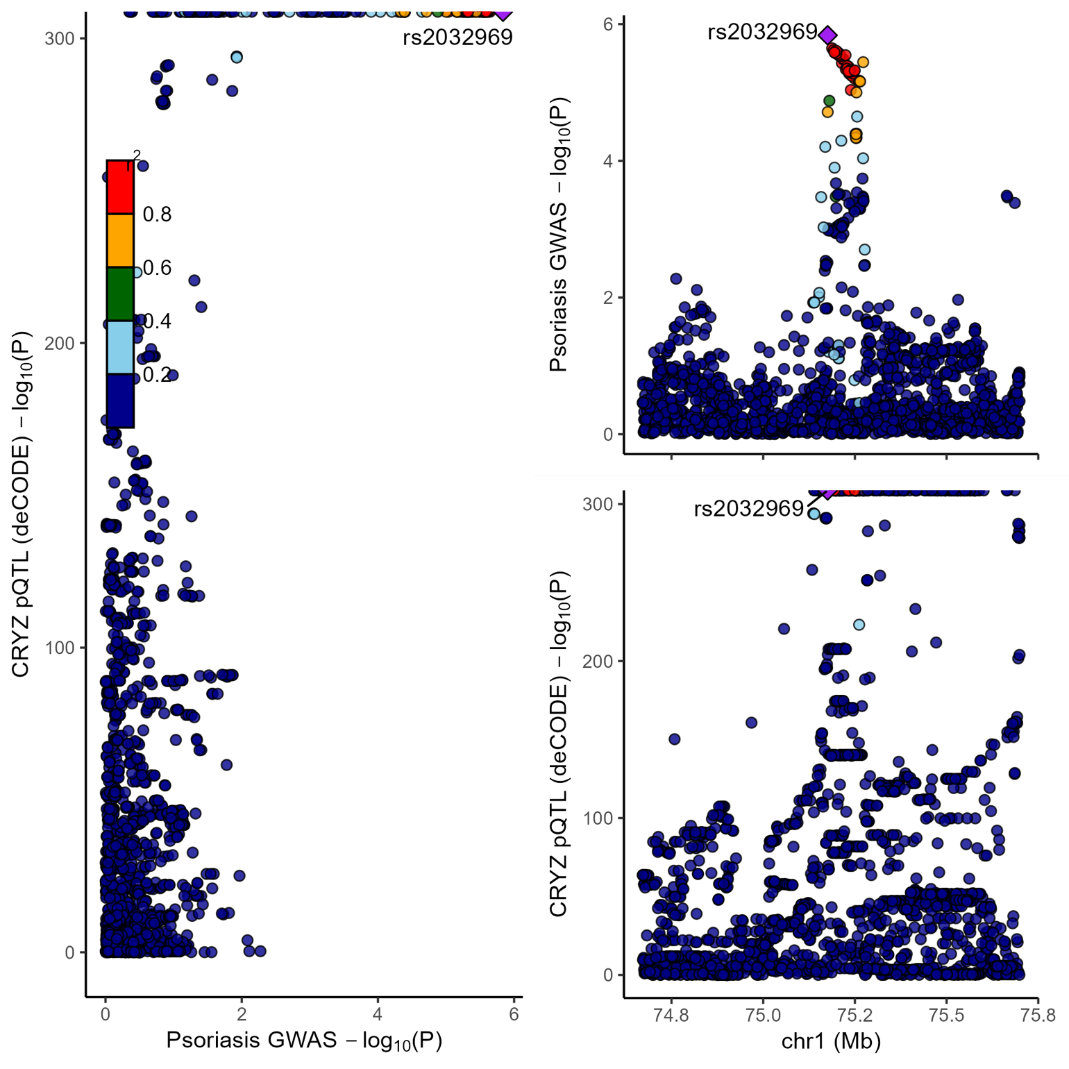

**Figure S23**. Locus Compare plot. Left-hand panel represents psoriasis GWAS (y-axis) and blood pQTL for XCL1 (x-axis) -log p values for the gene variants at the XCL1 locus. Right-hand panels represent association data with the -log p values on the y-axis for the pQTL (XCL1) (upper panel) and psoriasis GWAS (lower panel) according to genomic coordinates (x-axis) spanning 1Mb centered on XCL1.

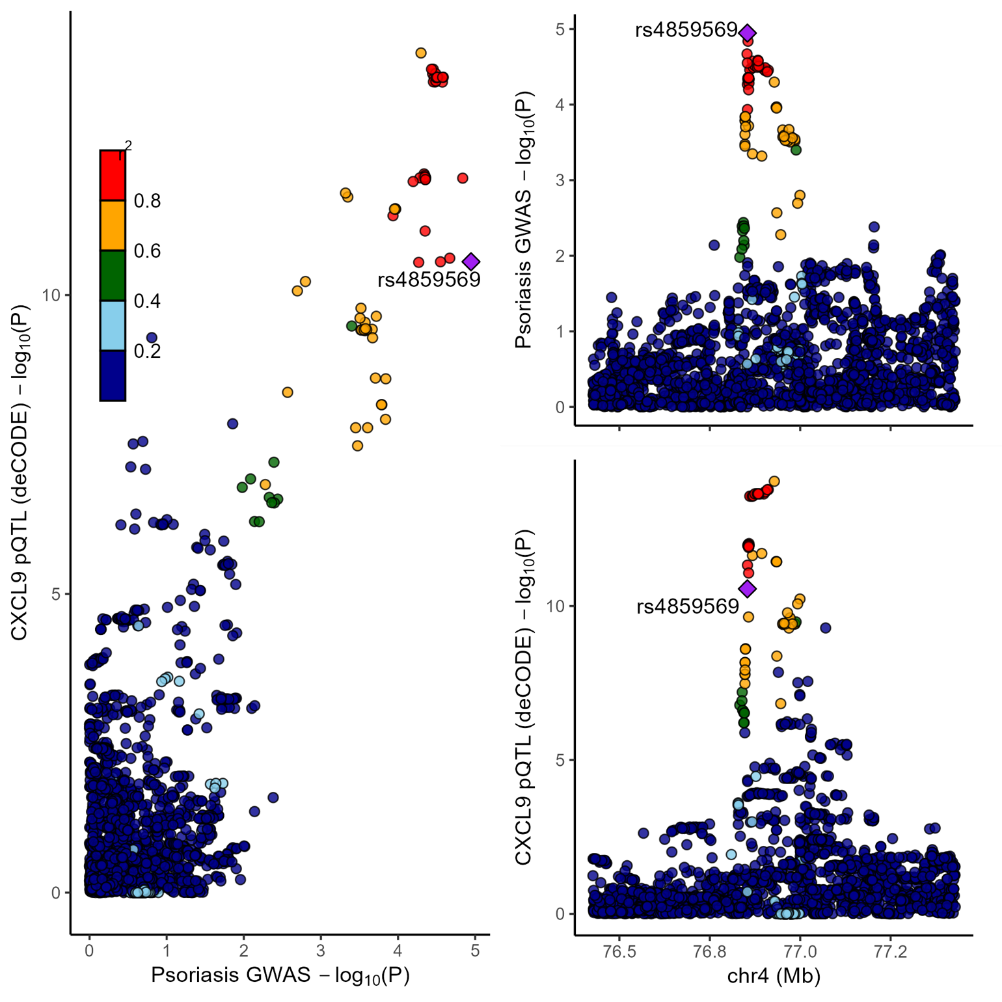

**Figure S24**. Locus Compare plot. Left-hand panel represents psoriasis GWAS (y-axis) and blood pQTL for XCL1 (x-axis) -log p values for the gene variants at the XCL1 locus. Right-hand panels represent association data with the -log p values on the y-axis for the pQTL (XCL1) (upper panel) and psoriasis GWAS (lower panel) according to genomic coordinates (x-axis) spanning 1Mb centered on XCL1.

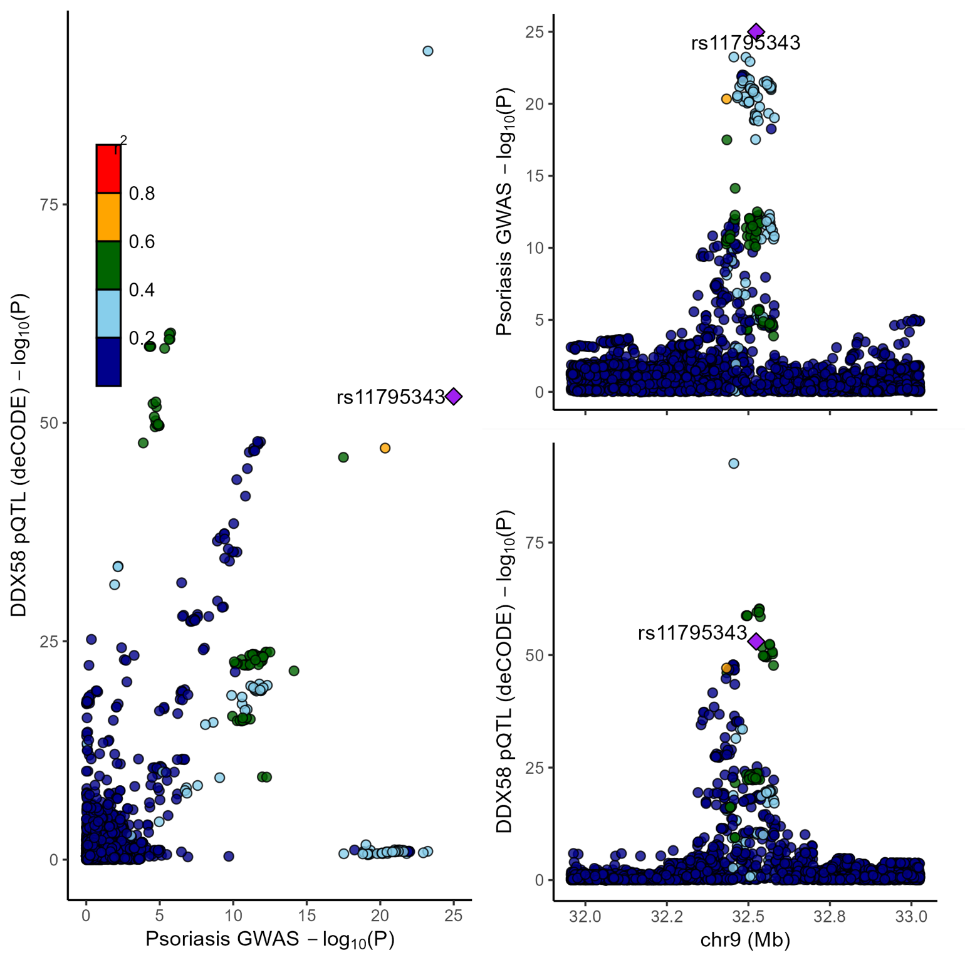

**Figure S25**. Locus Compare plot. Left-hand panel represents psoriasis GWAS (y-axis) and blood pQTL for DDX58 (x-axis) -log p values for the gene variants at the DDX58 locus. Right-hand panels represent association data with the -log p values on the y-axis for the pQTL (DDX58) (upper panel) and psoriasis GWAS (lower panel) according to genomic coordinates (x-axis) spanning 1Mb centered on DDX58.

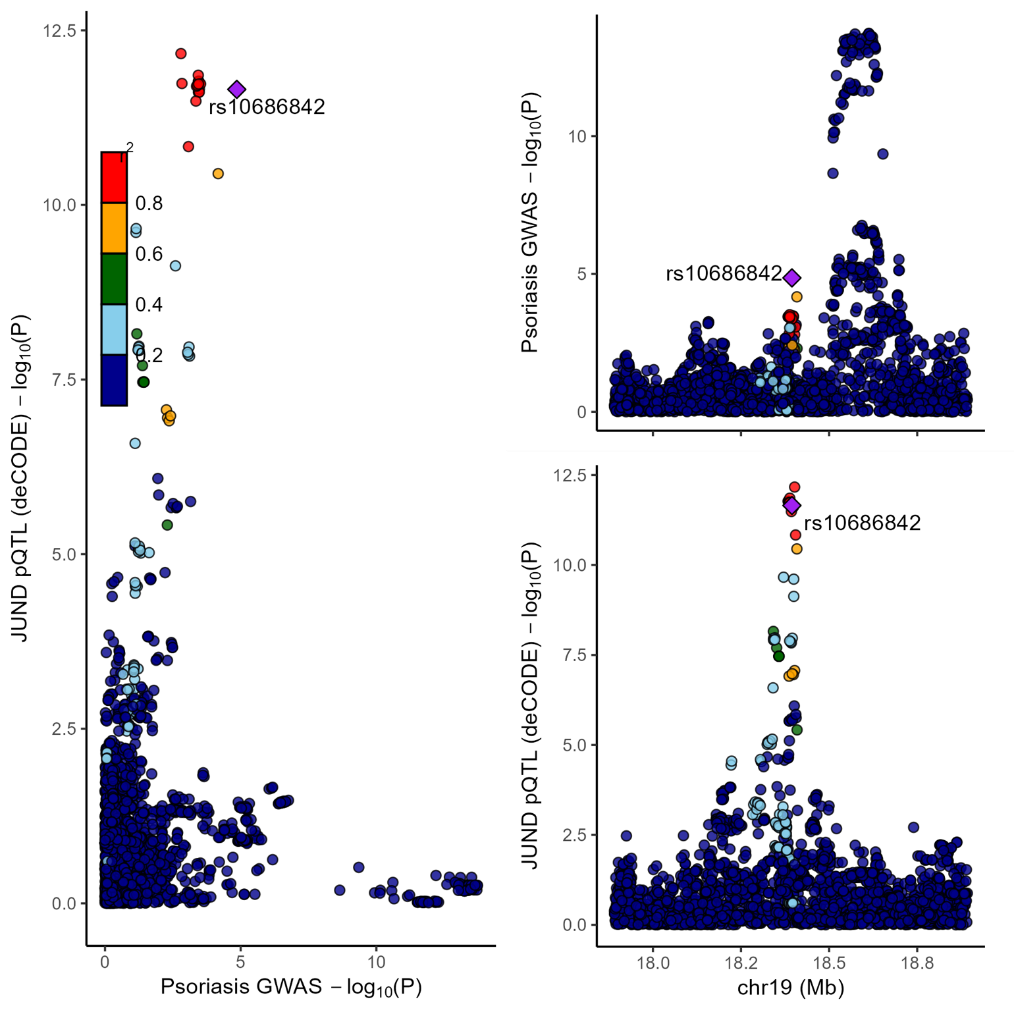

**Figure S26**. Locus Compare plot. Left-hand panel represents psoriasis GWAS (y-axis) and blood pQTL for JUND (x-axis) -log p values for the gene variants at the JUND locus. Right-hand panels represent association data with the -log p values on the y-axis for the pQTL (JUND) (upper panel) and psoriasis GWAS (lower panel) according to genomic coordinates (x-axis) spanning 1Mb centered on JUND.

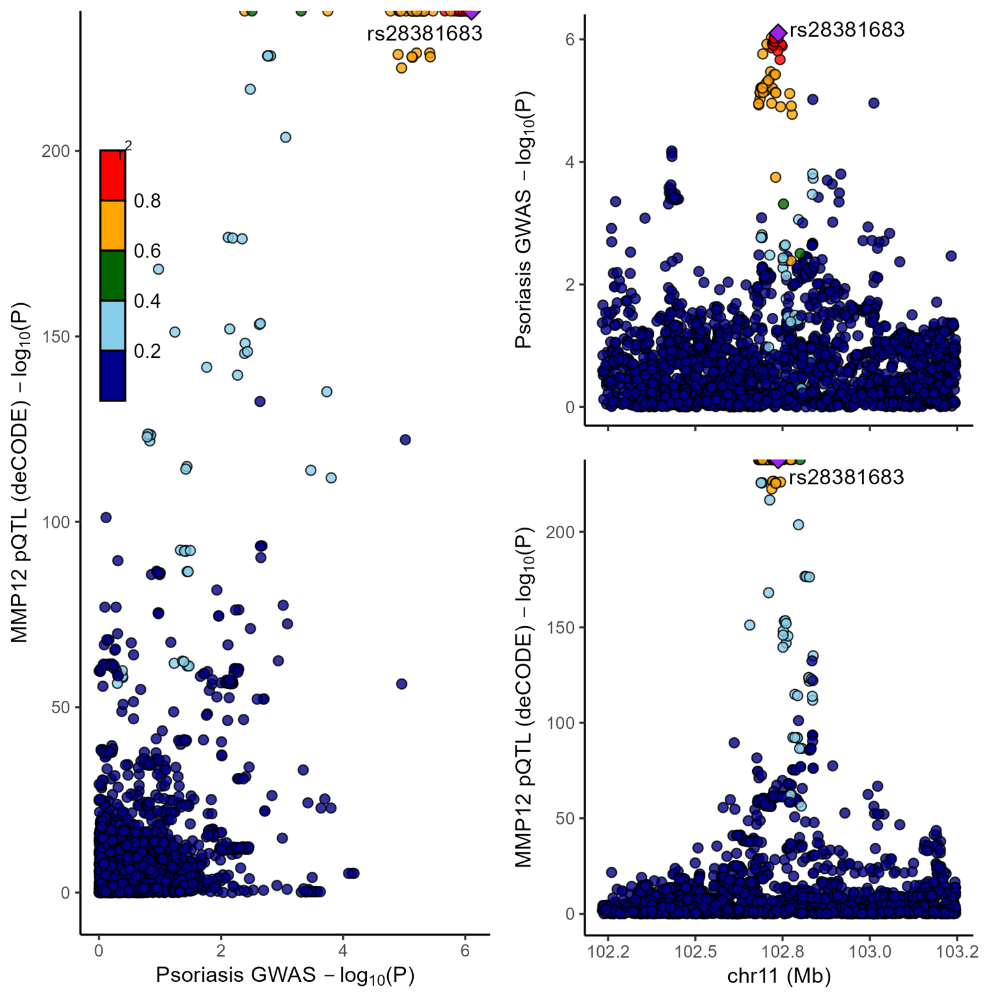

**Figure S27**. Locus Compare plot. Left-hand panel represents psoriasis GWAS (y-axis) and blood pQTL for MMP12 (x-axis) -log p values for the gene variants at the MMP12 locus. Right-hand panels represent association data with the -log p values on the y-axis for the pQTL (MMP12) (upper panel) and psoriasis GWAS (lower panel) according to genomic coordinates (x-axis) spanning 1Mb centered on MMP12.

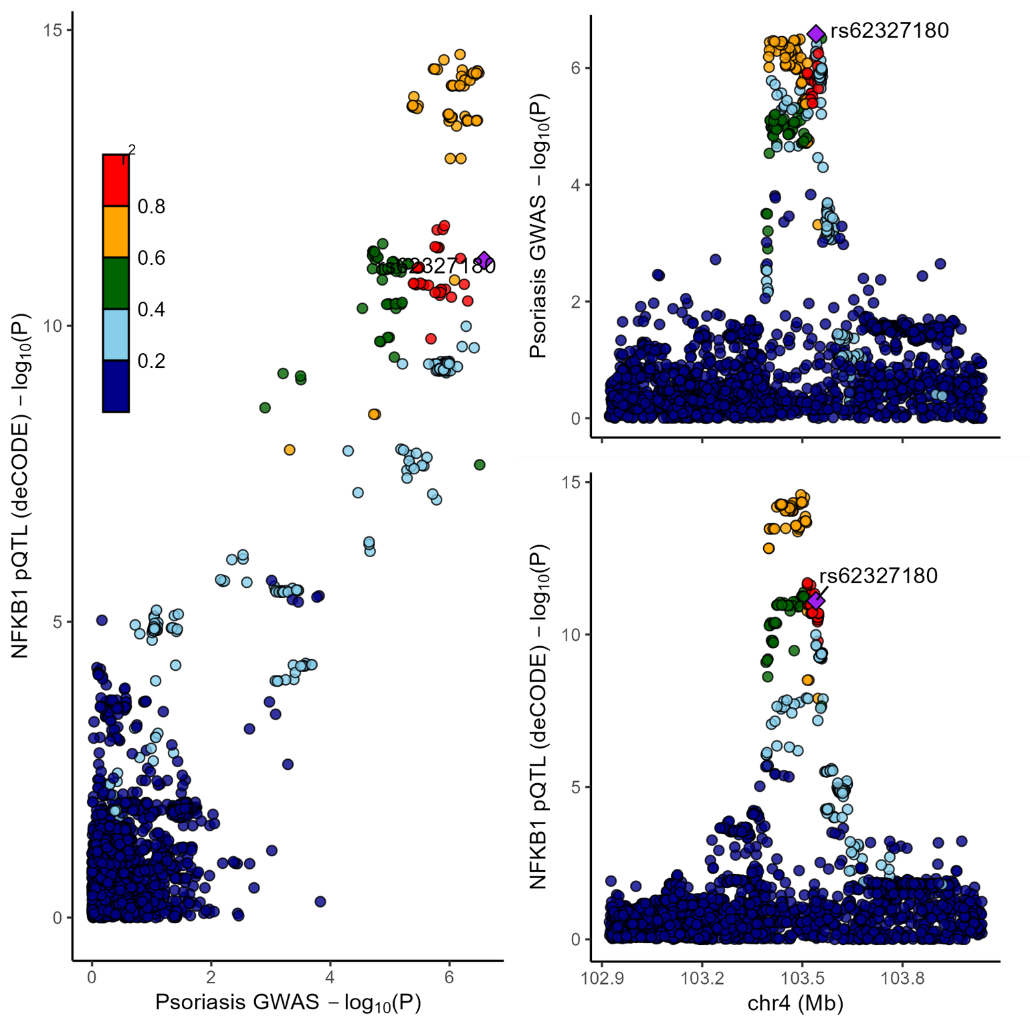

**Figure S28**. Locus Compare plot. Left-hand panel represents psoriasis GWAS (y-axis) and blood pQTL for NFKB1 (x-axis) -log p values for the gene variants at the NFKB1 locus. Right-hand panels represent association data with the -log p values on the y-axis for the pQTL (NFKB1) (upper panel) and psoriasis GWAS (lower panel) according to genomic coordinates (x-axis) spanning 1Mb centered on NFKB1.

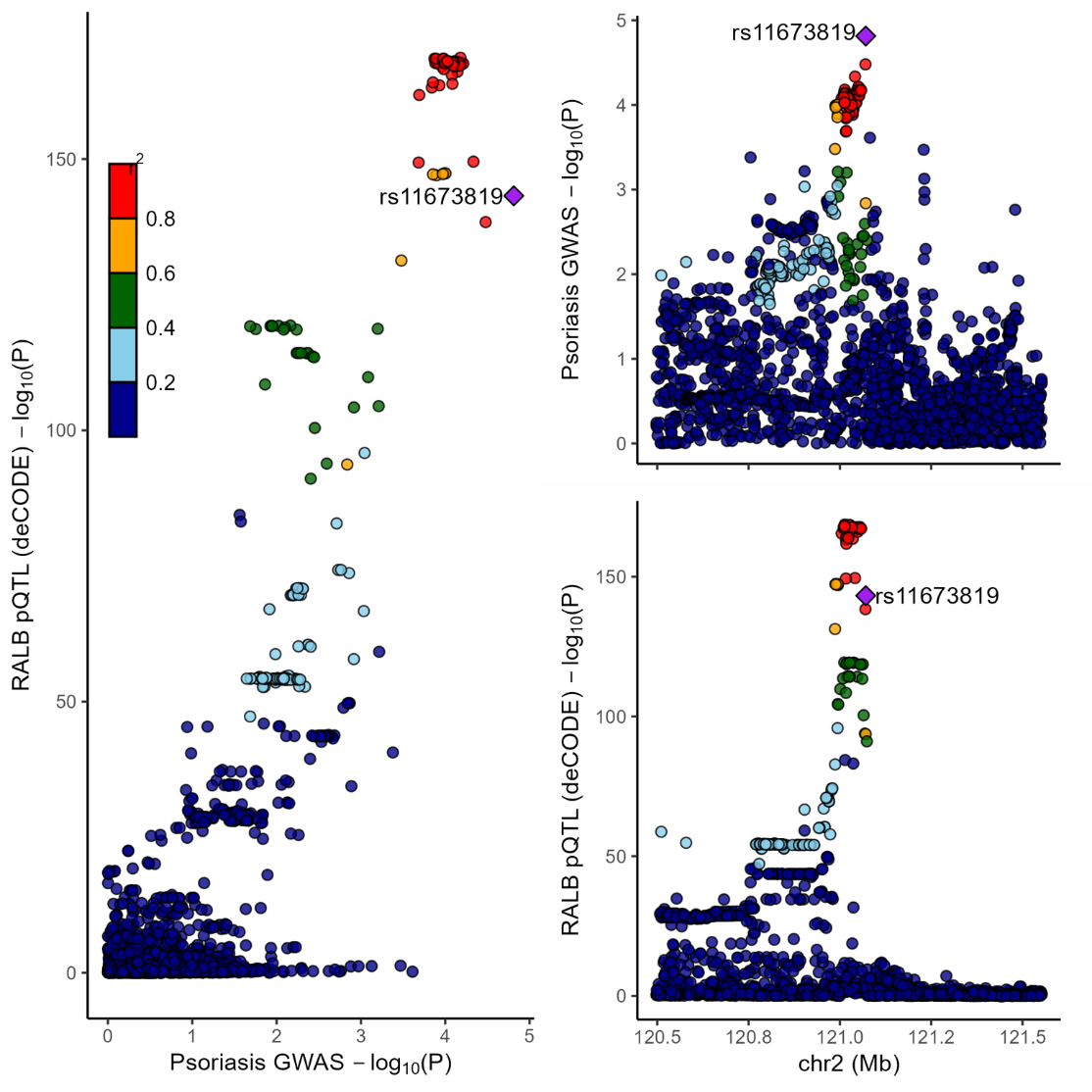

**Figure S29**. Locus Compare plot. Left-hand panel represents psoriasis GWAS (y-axis) and blood pQTL for RALB (x-axis) -log p values for the gene variants at the RALB locus. Right-hand panels represent association data with the -log p values on the y-axis for the pQTL (RALB) (upper panel) and psoriasis GWAS (lower panel) according to genomic coordinates (x-axis) spanning 1Mb centered on RALB.

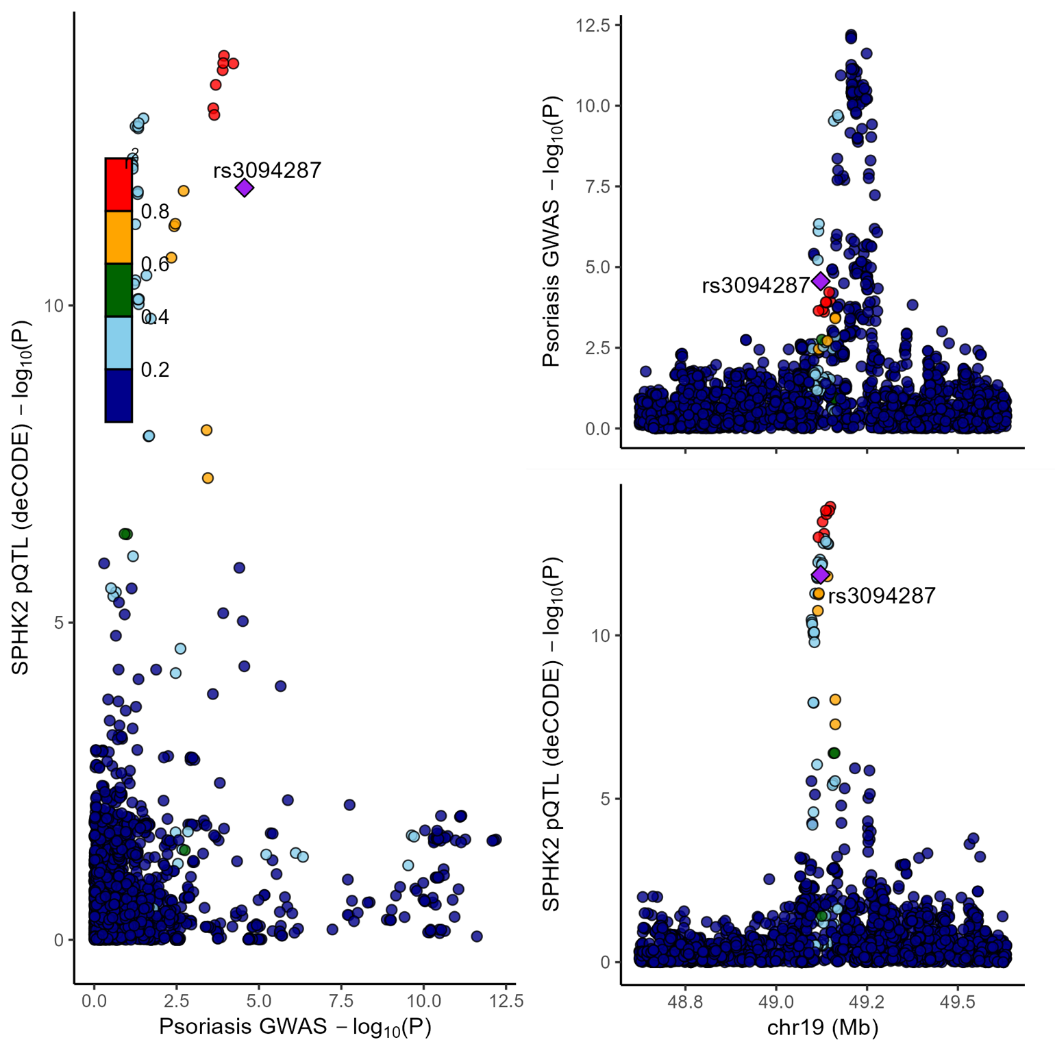

**Figure S30.** Locus Compare plot. Left-hand panel represents psoriasis GWAS (y-axis) and blood pQTL for SPHK2 (x-axis) -log p values for the gene variants at the SPHK2 locus. Right-hand panels represent association data with the -log p values on the y-axis for the pQTL (SPHK2) (upper panel) and psoriasis GWAS (lower panel) according to genomic coordinates (x-axis) spanning 1Mb centered on SPHK2.

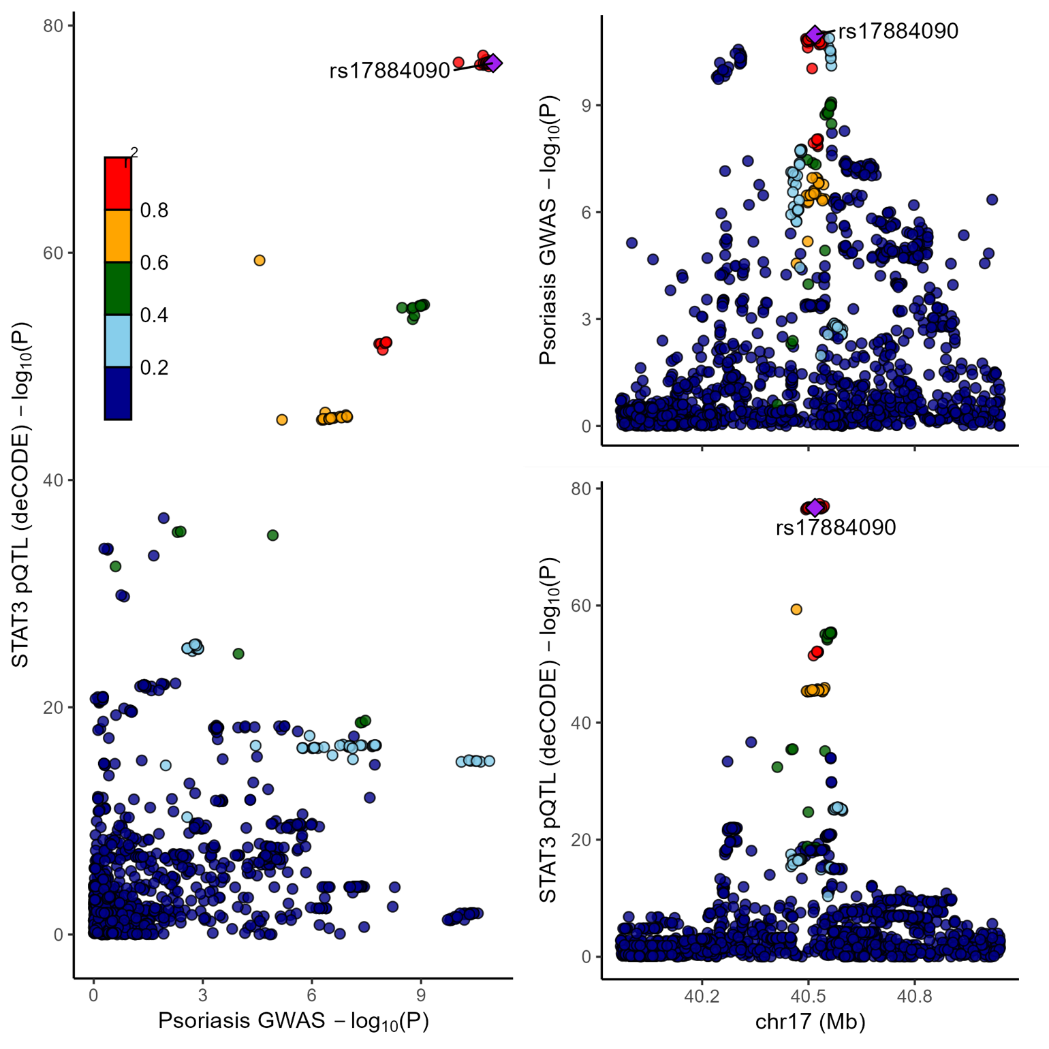

**Figure S31.** Locus Compare plot. Left-hand panel represents psoriasis GWAS (y-axis) and blood pQTL for STAT3 (x-axis) -log p values for the gene variants at the STAT3 locus. Right-hand panels represent association data with the -log p values on the y-axis for the pQTL (STAT3) (upper panel) and psoriasis GWAS (lower panel) according to genomic coordinates (x-axis) spanning 1Mb centered on STAT3.

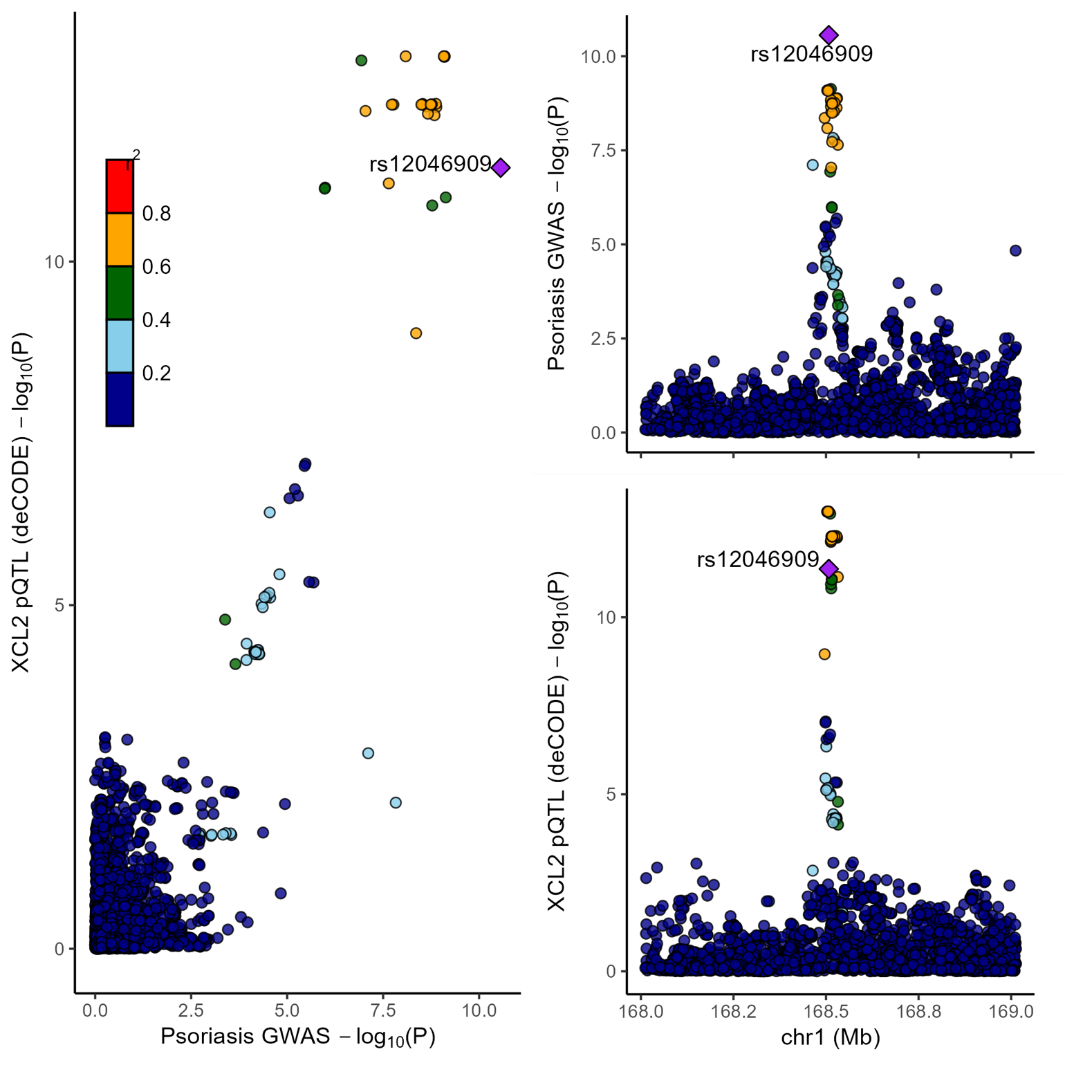

**Figure S32.** Locus Compare plot. Left-hand panel represents psoriasis GWAS (y-axis) and blood pQTL for XCL2 (x-axis) -log p values for the gene variants at the XCL2 locus. Right-hand panels represent association data with the -log p values on the y-axis for the pQTL (XCL2) (upper panel) and psoriasis GWAS (lower panel) according to genomic coordinates (x-axis) spanning 1Mb centered on XCL2.

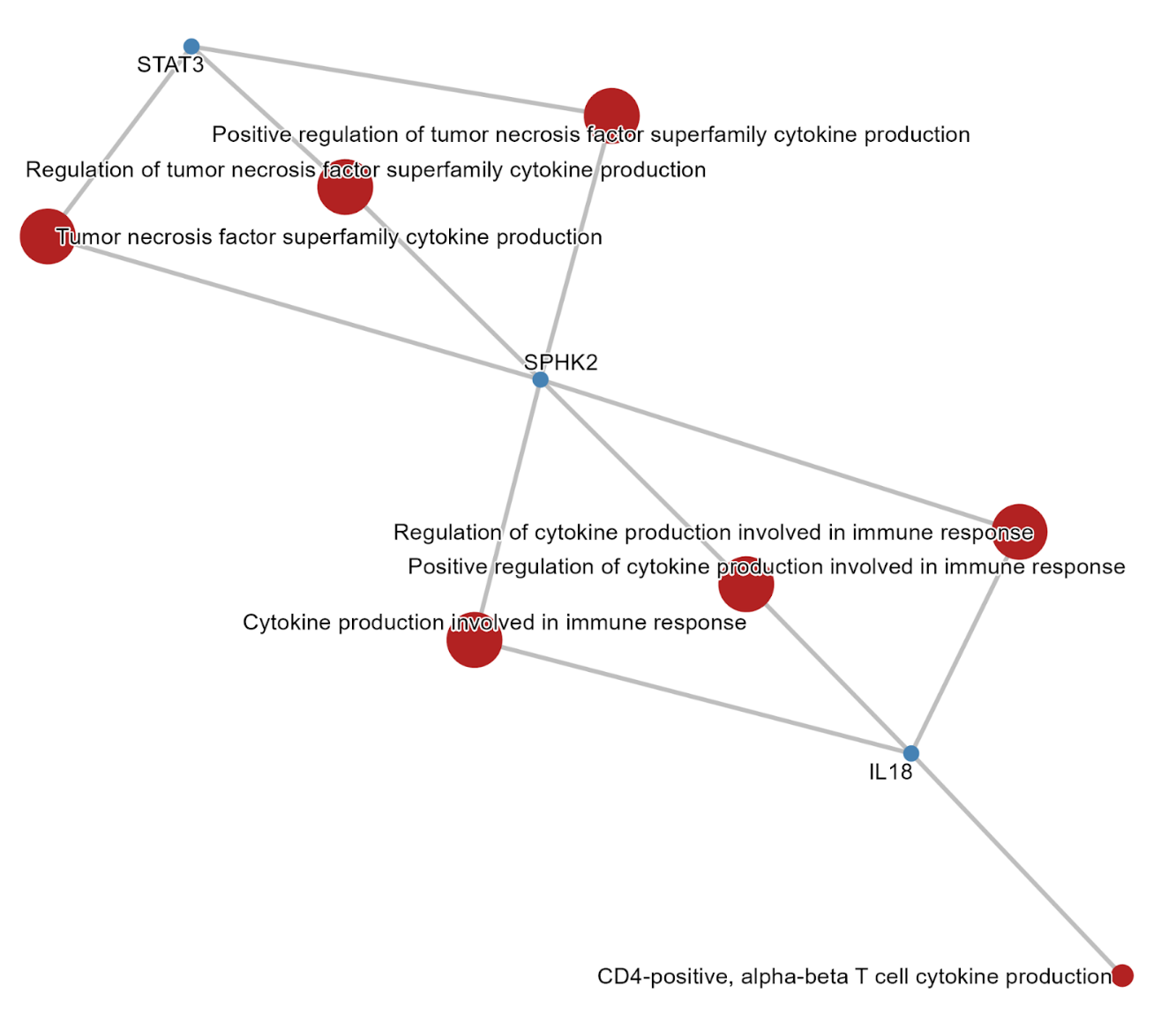

**Figure S33**. Gene Ontology (GO) network of enriched cytokine-related pathways among Tier 1 proteins.
Network plot showing significantly enriched GO biological processes (red nodes) linked to prioritised proteins (blue nodes). Node size reflects enrichment significance, and edges indicate gene–pathway associations.

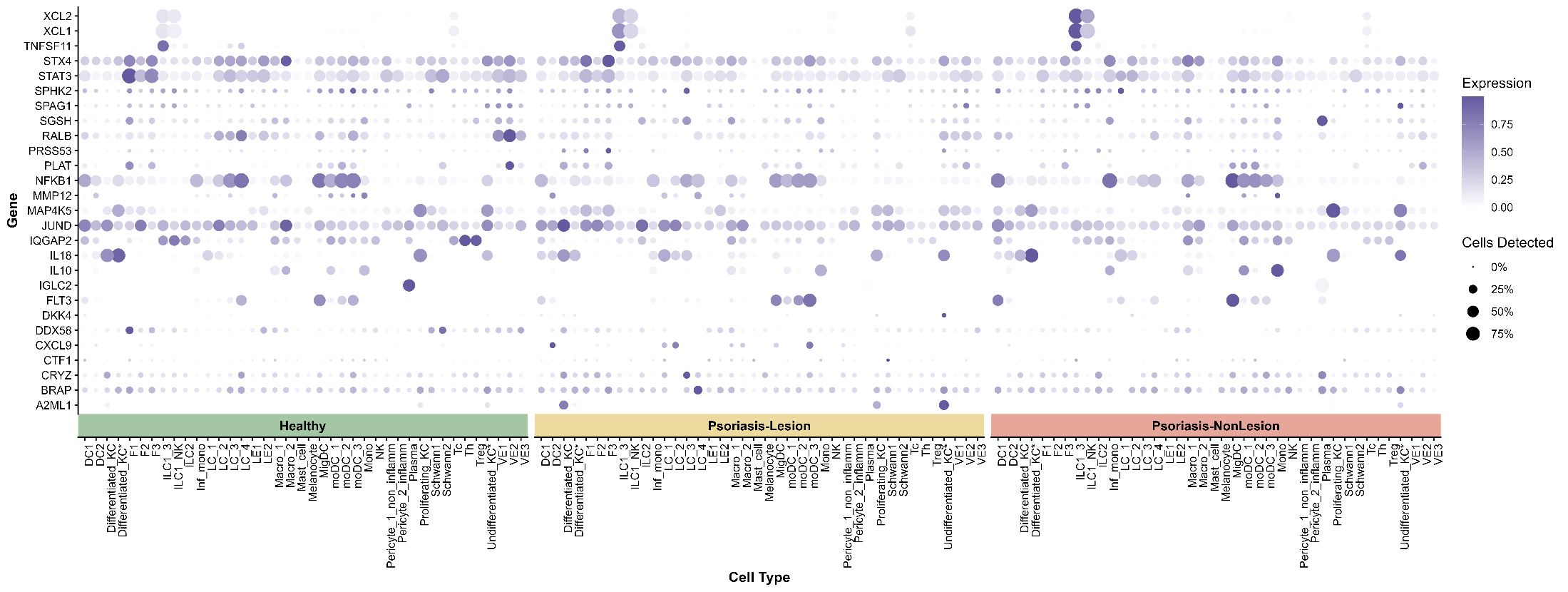

**Figure S34**. Cell-type–specific expression of prioritised genes across healthy, psoriatic lesional, and non-lesional skin. Dot plot showing the expression patterns of genes encoding prioritised proteins across major epithelial, stromal, and immune cell populations. Columns represent annotated cell types grouped by condition (Healthy, Psoriasis Lesional, Psoriasis Non-lesional), and rows represent genes. Dot size indicates the percentage of cells expressing the gene (>0 expression), and colour intensity represents scaled average expression levels.
